# Effects of the maternal and fetal proteome on birth weight: a Mendelian randomization analysis

**DOI:** 10.1101/2023.10.20.23297135

**Authors:** Nancy McBride, Alba Fernández-Sanlés, Marwa Al Arab, Tom A. Bond, Jie Zheng, Maria C. Magnus, Elizabeth C. Corfield, Gemma L Clayton, Liang-Dar Hwang, Robin N. Beaumont, David M. Evans, Rachel M. Freathy, Tom R. Gaunt, Deborah A Lawlor, Maria Carolina Borges

## Abstract

**Background:** Fetal growth is an important indicator of survival, regulated by maternal and fetal genetic and environmental factors. However, little is known about the underlying molecular mechanisms. Proteins play a major role in a wide range of biological processes and could provide key insights into maternal and fetal molecular mechanisms regulating fetal growth.

**Method:** We used intergenerational two-sample Mendelian randomization to explore the effects of 1,139 maternal and fetal genetically-instrumented plasma proteins on birth weight. We used genome-wide association summary data from the Early Growth Genetics (EGG) consortium (n=406,063 with maternal and/or fetal genotype), with independent replication in the Norwegian Mother, Father and Child Cohort Study (MoBa; n=74,932 mothers and n=62,108 offspring). Maternal and fetal data were adjusted for the correlation between fetal and maternal genotype, to distinguish their independent genetic effects.

**Results:** We found that higher genetically-predicted maternal levels of NEC1 increased birth weight (mean-difference: 12g (95% CI [6g, 18g]) per 1 standard deviation protein level) as did PRS57 (20g [10g, 31g]) and ULK3 (140g [81g, 199g]). Higher maternal levels of Galectin_4 decreased birth weight (-206g [-299g, -113g]). In contrast, in the offspring, higher genetically-predicted offspring levels of NEC1 decreased birth weight (-10g [-16g, -5g]), alongside sLeptin_R (-8g [-12g, -4g]), and UBS3B (-78g [-116g, -41g]). Higher fetal levels of Galectin_4 increased birth weight (174g [89g, 258g]). We replicated these results in MoBa, and found supportive evidence for shared causal variants from genetic colocalization analyses and protein-protein network associations.

**Conclusions:** We find strong evidence for causal effects, sometimes in opposing directions, of maternal and fetal genetically-instrumented proteins on birth weight. These provide new insights into maternal and fetal molecular mechanisms regulating fetal growth, involving glucose metabolism, energy balance, and vascular function that could be used to identify new intervention targets to reduce the risk of fetal growth disorders, and their associated adverse maternal and fetal outcomes.

## Introduction

Birth weight is a valuable and widely used measure of fetal growth. Healthy fetal growth is essential to minimise adverse perinatal health outcomes, including miscarriage, stillbirth, preterm birth and associated neonatal and infant morbidity and mortality (1–7). Large-scale genome-wide association studies (GWAS) have revealed hundreds of genomic regions independently associated with birth weight (2, 3, 8–10). In addition, these GWAS have uncovered a complex interplay between maternal and fetal genomes (8, 9), in which some genetic variants have maternal or fetal-specific effects on birth weight, whereas other genetic variants have both maternal and fetal effects, sometimes in opposite directions. Despite these advances in mapping fetal and maternal genetic variants, understanding the molecular mechanisms underpinning fetal growth remains a challenge (6).

Proteins play a major role in a wide range of biological processes (11). They are essential for cellular growth and repair, and could provide key insights into maternal and fetal molecular mechanisms regulating fetal growth (10). Mendelian Randomization (MR) is a method which uses genetic variants associated with exposures as instrumental variables, to test the effect of those exposures on human traits and diseases. The method aims to mitigate causal effects being biased due to confounding and reverse causation that may explain conventional observational associations. Previous MR studies have established causality of maternal modifiable factors on offspring birth weight, including higher maternal adiposity and higher maternal circulating glucose levels on higher birth weight, and smoking and higher blood pressure on lower birth weight (12) (13, 14) (5) (15). They have also provided novel insights, such as a potential effect of higher maternal glutamine levels on higher birth weight (5) and metabolically favourable adiposity on lower birth weight (16). As such, MR and proteomics can be integrated to explore maternal and fetal proteins regulating fetal growth by using genetic variants associated with protein levels (i.e., protein quantitative trait loci - pQTL) as instrumental variables. As most drug targets are proteins, identifying the effects of maternal circulating proteins on birth weight could identify targets for drug development that might prevent fetal growth restriction and over-growth, and their associated adverse pregnancy and perinatal outcomes.

The aim of this study was to identify causal effects of maternal and fetal proteins on birth weight to highlight underlying molecular mechanisms. To achieve that, we used two-sample MR, in which large-scale GWAS of plasma proteomics and birth weight were combined to examine the causal effect of plasma proteins on offspring birth weight.

## Methods

We performed a two-sample MR study to identify maternal and fetal effects of plasma proteins on offspring birthweight, examining both maternal and fetal genetic effects (n=406,063 with maternal and/or fetal genotype). For proteins passing multiple testing correction (FDR p-value < 5%), we tested whether results replicated in an independent sample (n=74,932 mothers and n=62,108 offspring) and investigated whether MR results could be confounded by linkage disequilibrium using genetic colocalization. For proteins which had discovery and replication evidence, and no evidence against colocalization, we used protein network associations to examine plausible pathways through which they could be influencing birth weight. **Figure 1** summarises the analytical workflow used in this paper.

**Figure 1.**
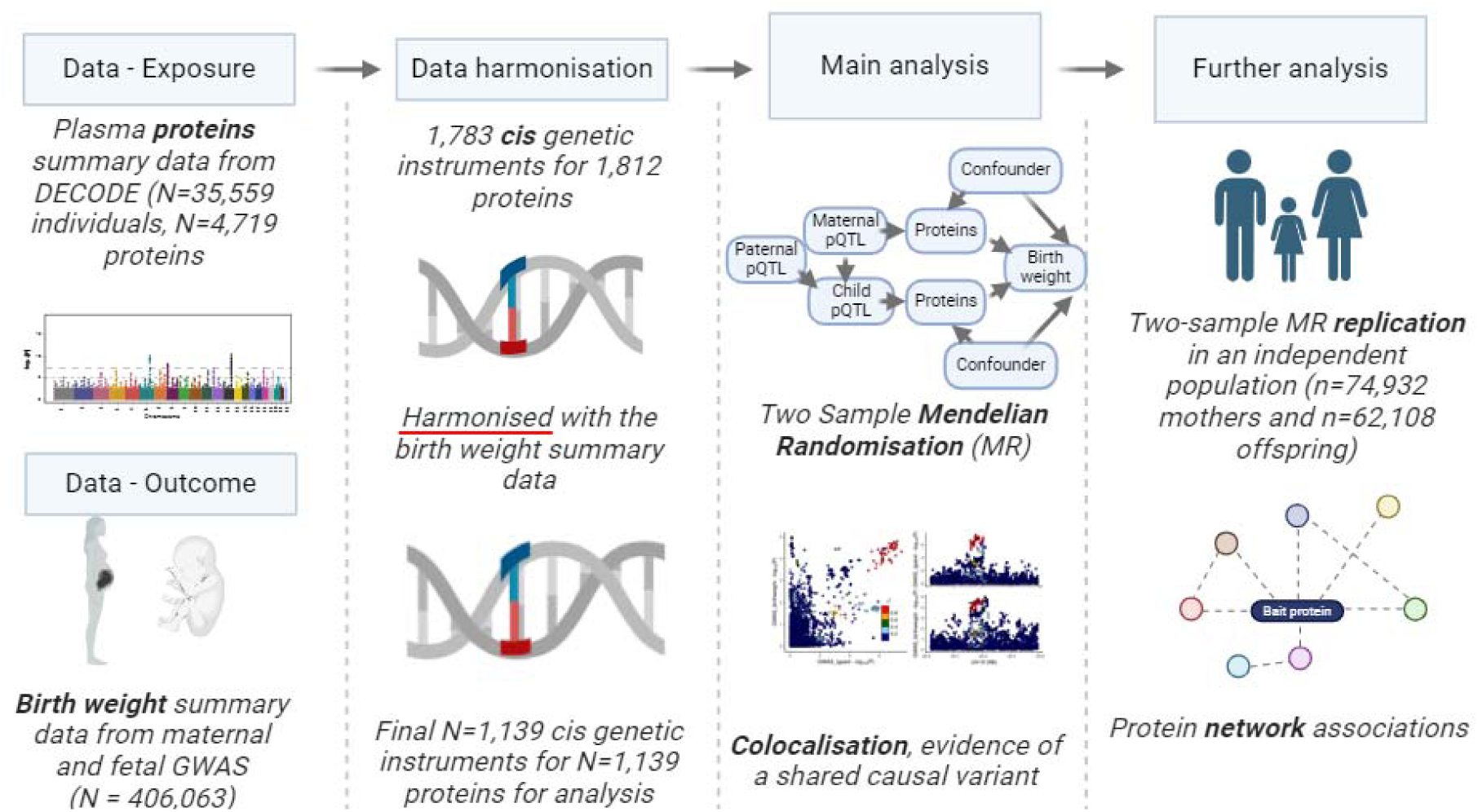
Schematic representation of the analyses conducted in this paper. Abbreviations: GWAS, genome wide association studies; pQTL, protein quantitative trait loci Selection of genetic instruments for plasma proteins We selected genetic instruments from the deCODE GWAS of plasma proteins, which provides data on 4,459 unique proteins, mapped to 13,514 pQTL. For our analyses, we excluded trans instruments (N=2,647 proteins with 6,402 trans pQTL) and secondary variants (N=5,238 secondary pQTL) retaining 1,783 cis pQTL defined as all those within ±1LJMb) for 1,812 proteins (*P*LJ<LJ1LJ×LJ10^−7^) **(Supplementary Figure 1)**

### Discovery sample

We used publicly available meta-analysed GWAS summary data on birth weight. These data were obtained from the 2019 Early Growth Genetics (EGG) consortium plus UK Biobank (UKBB) GWAS. This GWAS has publicly available estimates of maternal genetic associations adjusted for fetal genotype and fetal genetic associations adjusted for maternal genotype (see below ‘Adjustment for maternal/offspring genotype’).

The following exclusions were made prior to running the GWAS: i) twins and other multiple births, ii) offspring born earlier than 37 weeks of gestation, iii) extreme birth weight outliers (only individuals born between 2.2kg and 4.6kg were included in the maternal GWAS, and individuals born between 2.5 and 4.5kg were included in the offspring GWAS) and iv) babies born with congenital anomalies (for the maternal genotype only). Genetic associations with birth weight were reported by the GWAS in standard deviation (SD) units. We multiplied these by 551.36 (the SD of birth weight in the UKB) to obtain results in grams (g).

Overall, N=406,063 participants contributed to the models estimating the maternal and fetal genetic effects on birth weight that were adjusted, respectively, for fetal and maternal genotype. These included N=101,541 participants who reported their own birth weight and birth weight of their first child, N=195,815 participants with their own birth weight (fetal GWAS), and N=108,707 female participants with data on offspring birth weight (maternal GWAS)(8) (17).

### Replication study

For independent replication, we used genetic association data for birth weight from MoBa, a population-based pregnancy cohort study conducted by the Norwegian Institute of Public Health. Pregnant women and their partners were recruited from all over Norway from 1999-2008. The women consented to participate in 41% of the pregnancies. The cohort includes approximately N=114,500 children, N=95,200 mothers, and N=75,200 fathers (18). The current study uses version 12 of the quality-assured genetic data files released for research in January 2019. Birthweight was obtained from linked clinical records held in Medical Birth Registry of Norway (MBRN, a national health registry established in 1967 that contains information about all births in Norway, which is linked to the MoBa study using unique personal identification numbers). Questionnaire responses from mothers and fathers data were used to identify sex registered at birth, year of birth, birth weight, and multiple births (only available in the offspring). As in the discovery GWAS, multiple births and extremes of birth weight were excluded. Analyses were adjusted for principal components (PCs) and genotyping batch.

In MoBa, plasma blood samples were obtained from both parents during pregnancy, and from umbilical cord at birth (19). This project used MoBa genetic data that was quality controlled and imputed using the MoBaPsychGen pipeline, which has previously been described (20). Phasing and imputation were performed using the publicly available Haplotype Reference Consortium release 1.1 panel as a reference (21). The pipeline output consists of best-guess hard-call genotype data for 6,981,748 variants in 207,409 MoBa participants of European ancestry. GWAS in MoBa were run using *regenie* software (v 3.1.2) on the TSD server (22).

### Adjustment for maternal/offspring genotype

Birth weight is influenced by both maternal and fetal genotype. Therefore, to distinguish maternal and fetal genetic effects, in the discovery GWAS a weighted linear model was used to estimate maternal genetic associations (adjusted for fetal genotype), and fetal genetic associations adjusted for maternal genotype (3). From this, genetically-instrumented maternal effects of proteins on birth weight (not mediated or biased due to horizontal pleiotropy via fetal effects) and genetically instrumented fetal effects of proteins on birth weight (not confounded by maternal genotype) can be estimated. For our discovery sample (EGG+UKB GWAS), we downloaded these adjusted estimates from the GWAS consortium website. For our replication sample (MoBa), we estimated adjusted associations using the weighted linear model implemented via the DONUTS software package (51). As part of this, we used genetic covariance intercepts from LD score regression (LDSC) to account for overlap between maternal and fetal GWAS sample (23). The summary maternal adjusted for fetal, and fetal adjusted for maternal results were then used in our replication two-sample MR.

### Two sample MR

We conducted MR analyses to study the causal effects of circulating maternal and fetal proteins on birth weight. All MR analyses were performed using the *TwoSampleMR* R package version 0.5.6 (26). We used the Wald ratio estimator for analyses as only one pQTL was available per protein. Prior to MR analysis, genetic association data for proteins and birth weight were harmonized so that the effect of each genetic variant on the exposure and outcome were relative to the same effect allele. In some instances, data could not be harmonised (due to being palindromic SNPs, A/T or G/C, with allele frequencies close to 50%), and these pQTL and their corresponding proteins were removed from further analyses (**Supplementary Table 2)**. After excluding pQTL that failed harmonisation, we retained data for 1,139 proteins instrumented by the same number of *cis* pQTL (**Figure 1** and **Supplementary Figure 1**). We then analysed maternal and fetal data separately. MR results with a FDR-adjusted p-value ≤0.05 were carried forward to the replication MR and subsequent analyses.

We sought replication of the discovery MR results using genetic association data for birth weight from MoBa participants. There is no overlap between the discovery and replication participants. The same procedures for data harmonisation and two-sample MR analyses described in the discovery stage were used in the replication analyses. To report MR results as the mean difference in birth weight (g) per 1 SD increase in protein level, we multiplied MR effect estimates and standard error by 565.63 (g), which corresponds to 1 SD of birth weight in MoBa. We then conducted a fixed effects meta-analysis of estimates from discovery and replication samples using the *meta* R package (version 6.5-0). Our criteria for replication were: i) directionally consistent results between discovery and replication, ii) associations with a p-value threshold <0.05 in the meta-analysis, as commonly used in GWAS, and reflecting the relatively modest size of the replication sample in comparison to the main analysis sample and (iii) there was no strong evidence supporting heterogeneity between discovery and replication MR estimates (Cochrane’s Q p-value > 0.05).

### Genetic colocalization

We undertook genetic colocalization analyses using the *coloc* R package (24) to investigate whether our MR findings were compatible with a shared causal variant or confounded by linkage disequilibrium (LD) (24). This could happen if a selected pQTL is in LD with another genetic variant influencing birth weight via another biological mechanism. *Coloc* computes all possible configurations of causal variants for each of two traits and uses a Bayesian approach to calculate support for each causal model (H_0_: no association; H_1_: association with protein only; H_2_: association with birth weight only; H_3_: both traits owing to distinct causal variants; H_4_: both traits owing to a shared causal variant). We restricted the analyses to a ±500kb window around the pQTL, and assumed a prior probability that any pQTL in the region is associated with the protein only (p_1_) of 1 × 10^-4^, birth weight only (p_2_) of 1 × 10^-4^, or both traits (p_12_) 1 × 10^-5^. We considered a posterior probability of association (PPA) for H_4_ > 0.70 to provide strong evidence for colocalization, while a PPA for H_3_ > 0.5 was considered as evidence against colocalization. To facilitate visualisation of genomic signals in each region, we generated stacked recombination plots using the *locuscompareR* R package (http://locuscompare.com/) (25).

Where available, we extracted genetic association data for the genomic regions selected for colocalization analyses for plasma proteins and birth weight using the IEU Open GWAS (https://gwas.mrcieu.ac.uk/). The summary statistics for the genetic associations were downloaded from the deCODE website https://www.decode.com/summarydata/.

Fetal expression lookup

One of the assumptions of MR is that the genetic instruments are statistically strongly associated with the phenotype in the population to which inference would be made (i.e. the relevance assumption). We were unable to determine whether our genetic instruments associated with fetal (in utero) circulating proteins, due to the absence of large-scale datasets with both offspring genetic data and cord-blood measures of circulating proteins. Instead, we explored whether the proteins with evidence from discovery and replication were expressed in fetal tissue. We used the single-cell transcriptomics data from the human cell atlas of fetal gene expression (26). The atlas was developed from 121 human fetal samples (post-conceptional age = 72-129 days) from 15 organs (eye, heart, intestine, kidney, liver, lung, muscle, pancreas, placenta, spleen, stomach, thymus, adrenal, cerebellum and cerebrum), profiling 4 million single cells using a sci-RNA-seq3 protocol. The authors used a clustering approach building on a form of principal components analysis (uniform manifold approximation and projection - UMAP) to generate a resource where fetal gene expression can be mapped across different organs, tissues and cell types (https://descartes.brotmanbaty.org/). We used this resource to check that our proteins of interest were expressed in fetal tissue, as a further line of evidence.

### Protein networks

We explored information on protein-protein associations (referred to as interactions in genetic research) from the STRING database (https://string-db.org/cgi) to gain insights into presumed biological processes underlying our findings. STRING collates information from all known protein-protein interactions databases and provides a visual network of predicted associations for a particular group of proteins, whereby the proteins are represented as ‘nodes’ and the lines which join them are the predicted functional associations. The colour of the line denotes the source of the interaction information (see figure legend) (27). We set the confidence score of a given association to be ‘high confidence’ and restricted to associations from curated databases (either KEGG, Reactome, BioCyc and Gene Ontology, as well as legacy datasets from PID and BioCarta), experimentally determined (where evidence comes from lab experiments), reported from gene fusions (from reported gene fusion events), gene co-occurrence (where gene families have similar occurrence patterns across genomes), co-expression (where gene are shown to be correlated in expression across a large number of datasets) and protein homology (where proteins may have similar structures).

## Results

### Identifying causal effects of maternal circulating proteins on offspring birth weight

We found evidence of causal effects of 5 maternal proteins (instrumented by 5 cis-pQTLs) on offspring birth weight that passed our 5% false discovery rate (FDR) adjusted p-value **(Figure 2).** We found evidence that higher NEC1 increased birth weight (mean-difference: 12g (95% CI: 6g, 18g) per 1 standard deviation protein level) as did PRS57 (20g (10g, 31g)) and ULK3 (140g (81g, 199g)). Higher maternal levels of Galectin_4 decreased birth weight (-206g (-299g, -113g)), as did PTN9 (-159g (-237g, -82g)). **(Figure 2)**.

**Figure 2.**
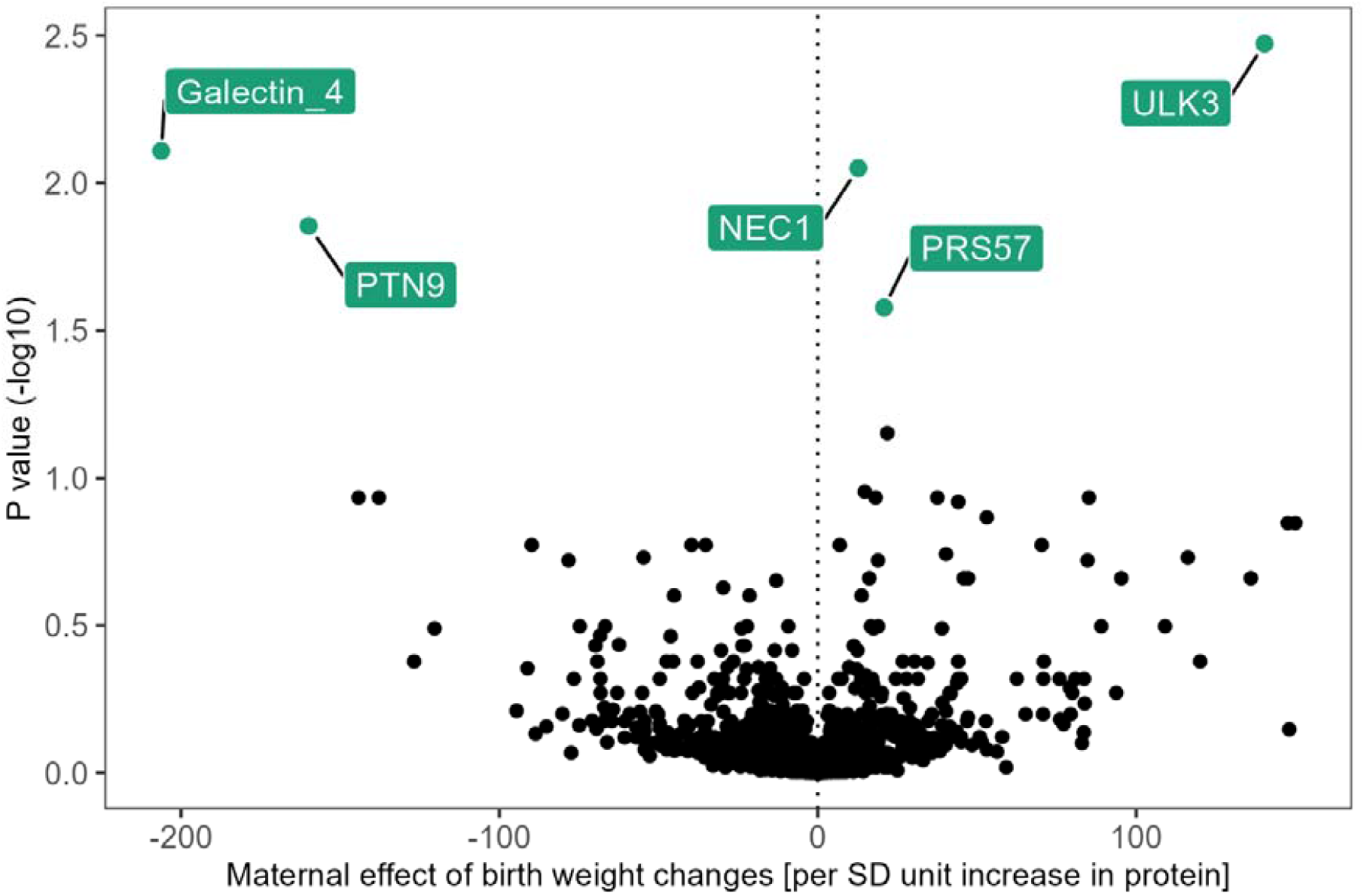
Two sample MR results using maternal cis pQTLs against offspring birth weight. FDR corrected p-values transformed to their -log10. Results plotted in green passed 5% FDR corrected p-value threshold. Abbreviations: SD, standard deviation

MR results for maternal proteins NEC_1, Galectin_4, ULK3 and PTN9 identified in discovery analyses met our replication criteria based on meta-analysed results from pooled discovery and replication. However, the replication result for PRS57 was not directionally consistent and there was evidence of high heterogeneity in the meta-analysis for PRS57, ULK3, PTN9 (**Figure 3** and **Supplementary Table 6 and 8**).

**Figure 3.**
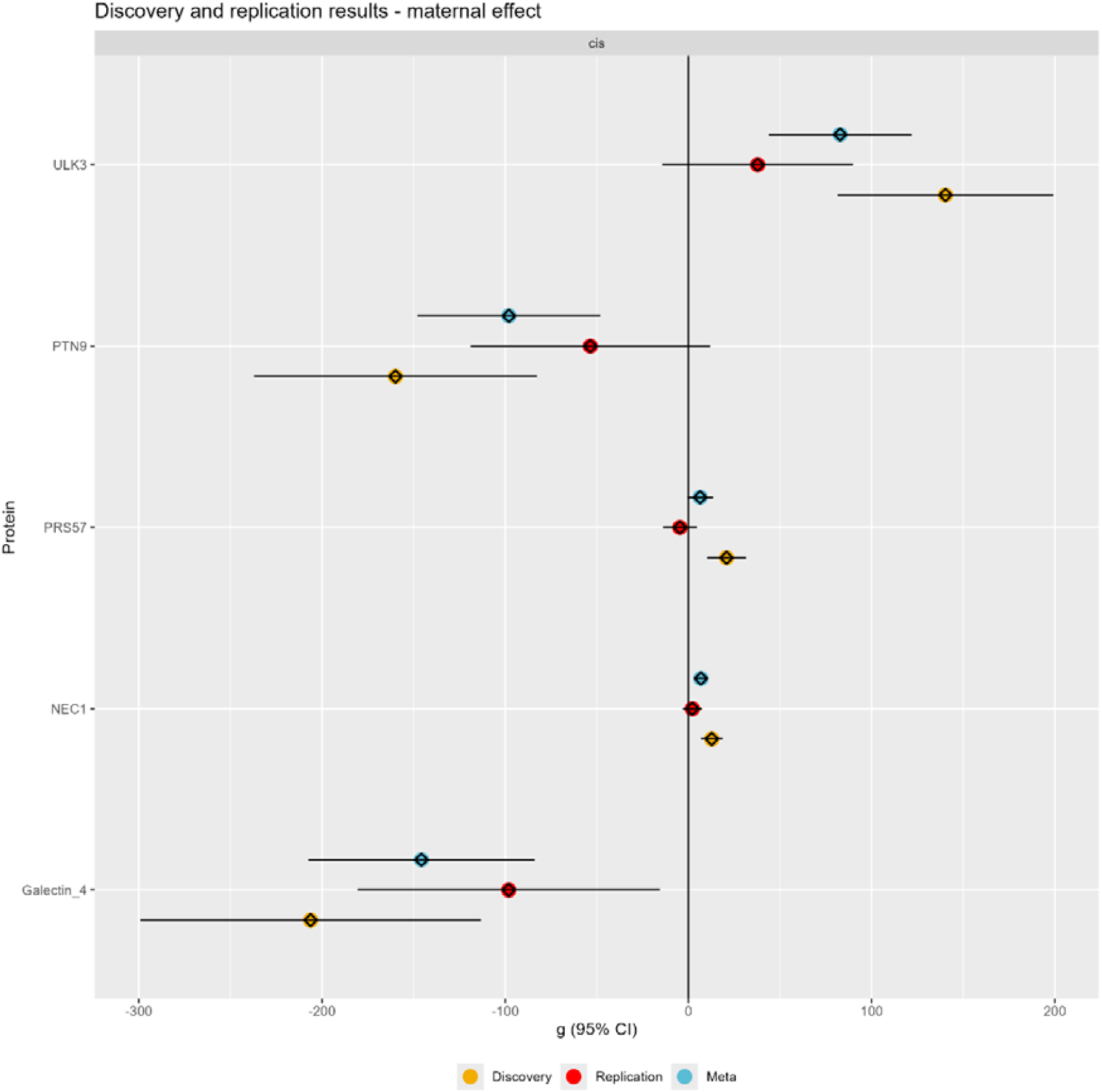
Mendelian randomization results estimating the effect of genetically instrumented maternal proteins (in standard deviation units) on offspring birth weight (in grams) in the discovery (orange dots N=210,248 mothers), replication (red dots; N=87,651 mothers) and meta-analysed discovery and replication data. Replication was tested for proteins passing FDR-corrected p-value <0.05 in the discovery data. Our criteria for replication were: i) directionally consistent results between discovery and replication, ii) associations with a p-value threshold <0.05 in the meta-analysis, as commonly used in GWAS, and reflecting the relatively modest size of the replication sample in comparison to the main analysis sample and (iii) there was no strong evidence supporting heterogeneity between discovery and replication MR estimates (Cochrane’s Q p-value > 0.05).

For maternal effects, we undertook colocalization analysis for all proteins which passed our FDR-adjusted p-value in the discovery analysis. For colocalization analyses, results are shown in **Table 1** as posterior probabilities (PP) of no association (H_0_), association with protein only (H_1_), birth weight only (H_2_), both traits owing to distinct causal variants (H_3_), or both traits owing to a shared causal variant (H_4_) (24). The colocalization analyses supported causal effects for maternal circulating NEC1, Galectin_4 and PRS57 with offspring birth weight, with high H_4_ of 0.82, 0.84, and 0.76 respectively **(Table 1)**. For PTN9 and ULK3, there was strong evidence that the association stemmed from distinct causal variants.

**Table 1.** Genetic colocalization results for maternal proteins with birth weight.

| Protein | $H_0$ | $H_1$ | $H_2$ | $H_3$ | $H_4$ |
| --- | --- | --- | --- | --- | --- |
| NEC1 | 0 | 0.05 | 0 | 0.13 | 0.82 |
| Galectin_4 | 0 | 0.06 | 0 | 0.10 | 0.84 |
| PTN9 | 0 | 0 | 0 | 0.99 | 0 |
| ULK3 | 0 | 0 | 0 | 0.93 | 0.06 |
| PRS57 | 0 | 0.21 | 0 | 0.03 | 0.76 |

Abbreviations – PP, posterior probabilities; H, hypothesis. Results are expressed as posterior probabilities (PP) of no association ( $H_0$ ), association with protein only ( $H_1$ ), birth weight only ( $H_2$ ), both traits owing to distinct causal variants ( $H_3$ ), or both traits owing to a shared causal variant ( $H_4$ ).

Maternal discovery and replication MR, and colocalization results of are summarised in **Table 2**. Together these provide strong evidence for maternal circulating proteins Galectin_4 and NEC1 being identified in discovery, replication, and not being biased due to confounded by LD (H_4_ 0.84 and 0.82, respectively). There was statistical support for PRS57 in the discovery and colocalization analysis (H_4_ 0.76), but not the replication. Whilst there was good replication for ULK3, colocalization results for ULK3 and PTN9 suggested results were being driven by distinct causal variants (H_3_ > 0.9).

**Table 2.** Summary of results from analyses exploring the effect of maternal proteins on offspring birth weight.

| Protein | Uniprot ID | Discovery MR direction of effect | Independently replicated | Colocalization |
| --- | --- | --- | --- | --- |
| NEC1 | P29120 | + | Yes | Yes |
| Galectin_4 | P56470 | - | Yes | Yes |
| PTN9 | Q99952 | - | Yes | No |
| ULK3 | Q6PHR2 | + | Yes | No |
| PRS57 | Q6UWY2 | + | No | Yes |

### Identifying causal effects of fetal circulating proteins on offspring birth weight

We found evidence of causal effects of 14 proteins on offspring birth weight. Higher genetically-predicted offspring levels of NEC1 decreased birth weight (-10g (-16g, -5g), alongside sLeptin_R (-8g (-12g, -4g)), UBS3B (-78g (-116g, -41g)), FUT5 (-19g (-29g, -9g)), IFN_a_b_R1 (-53g (-84g, -23g)), TWEAK (-24g, (-37g, -11g)) and MK13 (-69g (-108g, - 30g)). Higher fetal levels of Galectin_4 increased birth weight (174g (89g, 258g)), alongside ACADV (227g (142g, 311g)), PLCG1 (133g (72g, 193g)), BCMA (61g (27g, 96g)) Cystatin C (37g (16g, 58g), and FKB1B (46g (26g, 67g)) (**Figure 4**).

**Figure 4.**
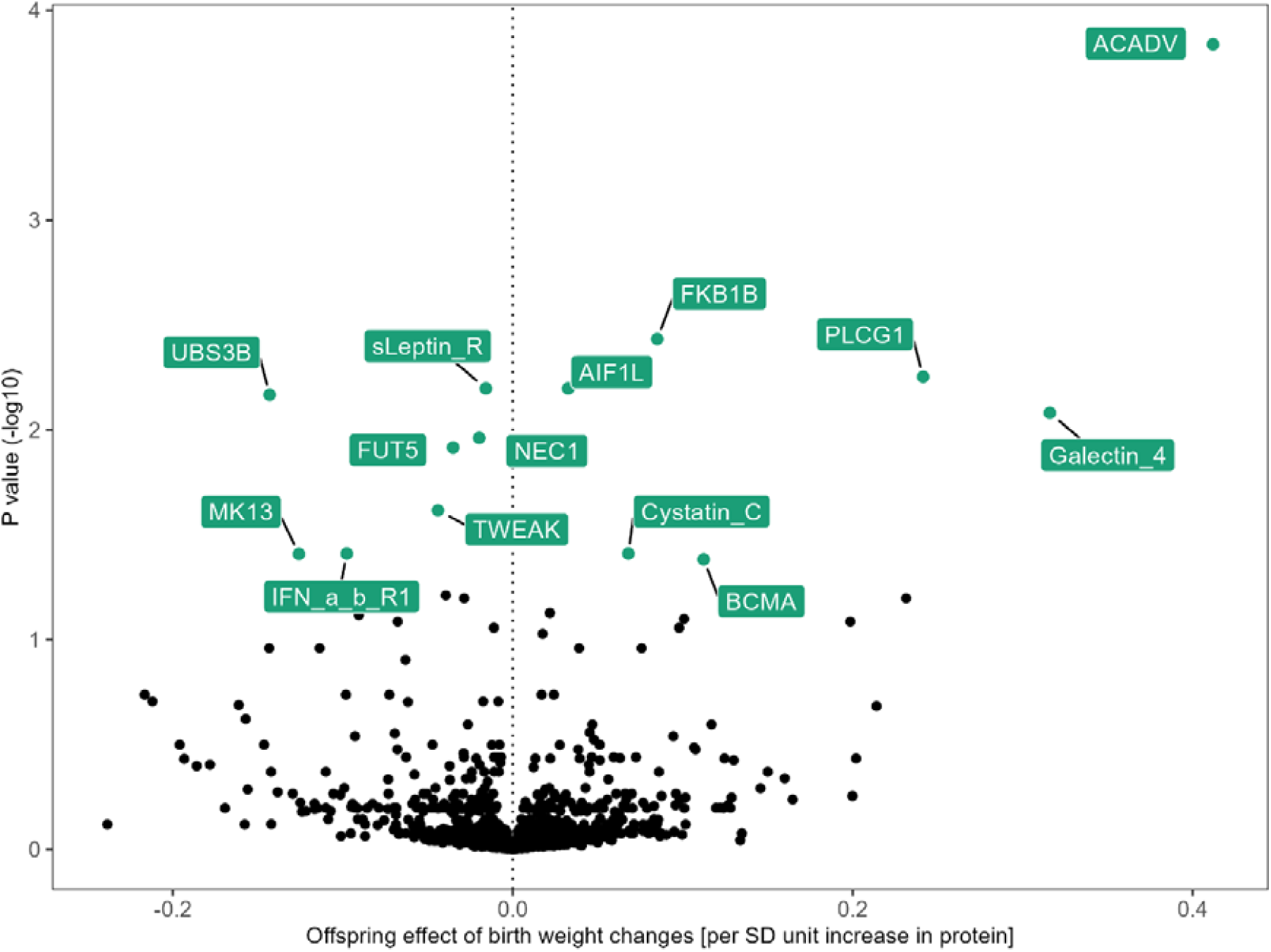
Fetal two sample MR results for cis pQTL. FDR corrected p-values transformed to their -log10. Results plotted in green passed FDR corrected p-value threshold. Abbreviations: SD, standard deviation

As in the maternal effects analysis, we conducted replication in MoBa and then ran a meta-analysis. We found meta-analysed MR results consistent with discovery for NEC1, Galectin_4, sLeptin_R, TWEAK, ACADV, UBS3B, BCMA (same direction of effect and with a meta-analysed FDR adjusted pvalue <0.05 and no evidence of between discovery and replication results heterogeneity). PLCG1, MK13, FUT5, FKB1B, and Cystatin_c did not replicate, and the replication sample estimate had the opposite direction of effect to the discovery sample (**Figure 5** and **Supplementary Table 7**). The pQTL for protein IFN_a_b_R1 was not in our replication data so could not be tested or meta-analysed (**Supplementary Table 9**).

**Figure 5.**
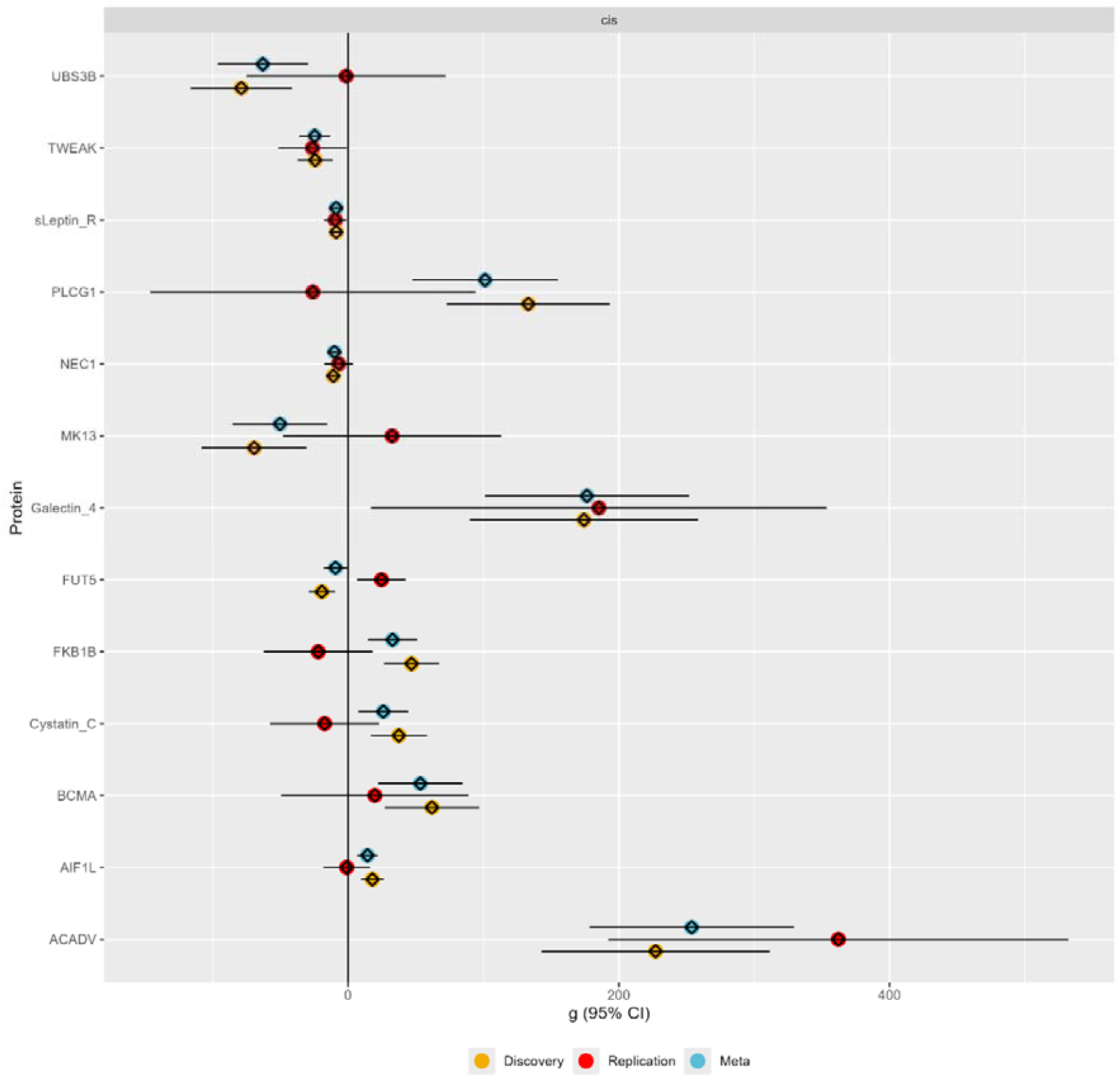
Mendelian randomization analyses estimating the effect of genetically-instrumented fetal proteins (in standard deviation units) on offspring birth weight (in grams) in the discovery (N=297,356 offspring, orange dots), replication (N=62,108 offspring, red dots) and meta-analyses (blue dots) of discovery and replication samples. Replication was tested for proteins passing FDR-corrected p-value <0.05 in the discovery data. Our criteria for replication were: i) directionally consistent results between discovery and replication, ii) associations with a p-value threshold <0.05 in the meta-analysis, as commonly used in GWAS, and reflecting the relatively modest size of the replication sample in comparison to the main analysis sample and (iii) there was no strong evidence supporting heterogeneity between discovery and replication MR estimates (Cochrane’s Q p-value > 0.05).

For fetal effects, we undertook colocalization analysis for all proteins which passed our FDR adjusted p-value in the discovery analysis. There was strong evidence of colocalization for NEC1, ACADV, PLCG1, FUT5, IFN_a_b_R1, AIF1L and UBS3B (all H_4_ > 0.75). There was moderate evidence of colocalization for sLeptin_R (H_4_ 0.65). There was evidence that the MR results for fetal Galectin_4, Cystatin-C, BCMA and MK13 might be explained by LD confounding (H_3_ > 0.5 or high H_1_) (**Table 3**).

**Table 3.** Genetic colocalization results for offspring proteins with birth weight.

| Protein | H<sub>0</sub> | H<sub>1</sub> | H<sub>2</sub> | H<sub>3</sub> | H<sub>4</sub> |
| --- | --- | --- | --- | --- | --- |
| NEC1 | 0 | 0.14 | 0 | 0.09 | 0.77 |
| Galectin_4 | 0 | 0.07 | 0 | 0.57 | 0.35 |
| ACADV | 0 | 0 | 0 | 0.16 | 0.83 |
| CystatinC | 0 | 0.42 | 0 | 0.08 | 0.49 |
| BCMA | 0 | 0.45 | 0 | 0.09 | 0.45 |
| TWEAK | 0 | 0 | 0.6 | 0.37 | 0 |
| FUT5 | 0 | 0.09 | 0 | 0.01 | 0.88 |
| FKB1B | 0 | 0 | 0 | 0.99 | 0 |
| PLCG1 | 0 | 0.03 | 0 | 0.09 | 0.86 |
| MK13 | 0 | 0.33 | 0 | 0.3 | 0.37 |
| IFN_a_b_R1 | 0 | 0.1 | 0 | 0 | 0.89 |
| AIF1L | 0 | 0.07 | 0 | 0 | 0.93 |
| UBS3B | 0 | 0.05 | 0 | 0.01 | 0.92 |
| sLeptin_R | 0 | 0.03 | 0 | 0.3 | 0.65 |

Abbreviations – PP, posterior probabilities; H, hypothesis. Results are expressed as posterior probabilities (PP) of no association (H<sub>0</sub>), association with protein only (H<sub>1</sub>), birth weight only (H<sub>2</sub>), both traits owing to distinct causal variants (H<sub>3</sub>), or both traits owing to a shared causal variant (H<sub>4</sub>).

Fetal discovery and replication MR, and colocalization results of are summarised in **Table 4**. Together these provide strong evidence for fetal circulating proteins.

| Protein | Uniprot ID | Discovery MR direction of effect | Independently replicated | Colocalization |
| --- | --- | --- | --- | --- |
| NEC1 | P29120 | - | Yes | Yes |
| Galectin_4 | O00214 | + | Yes | No |
| ACADV | P49748 | + | Yes | Yes |
| CystatinC | P01034 | + | No | No |
| BCMA | Q02223 | + | No | No |
| TWEAK | O43508 | - | Yes | No |
| FUT5 | Q11128 | - | No | Yes |
| FKB1B | Q96AY3 | + | No | No |
| PLCG1 | P19174 | + | No | Yes |
| MK13 | Q92910 | - | No | No |
| IFN_a_b_R1 | P14787 | - | NA | Yes |
| AIF1L | Q9H8A1 | + | No | Yes |
| UBS3B | Q8WUW0 | - | Yes | Yes |
| sLeptin_R | P48357 | - | Yes | Uncertain |

### Fetal expression lookup

As a sanity check of our fetal results, we were able to check whether these genes are expressed in fetal tissue using an atlas of human gene expression during development (https://descartes.brotmanbaty.org/). We wanted to see if tissue expression data supported expression of the proteins with evidence from discovery and replication or colocalization in fetal tissue. We found this was the case for genes encoding NEC1 (in pancreas), FUT5 (in lung and thymus), Galectin_4, UBS3B, sLeptin_R, AIF1L, PLCG1, IFN_a_b_R1 and ACADV (all expression all across multiple tissues) (**Supplementary Figures 20-29**).

### Protein networks of identified maternal and fetal proteins affecting birth weight

We undertook protein-protein association analyses for the proteins that replicated in the MR results, and had supporting colocalization results in either maternal (NEC1, Galectin_4) or fetal analyses (NEC1, ACADV, UBSH3B).

We found evidence that NEC1 (proprotein convertase 1, the same enzyme as PCSK1, as in the **Figure 6B** below) associates with proteins regulating glucose homeostasis, such as insulin (INS), and glucagon (GCG) - hormones produced by the pancreas to regulate blood sugar levels. NEC1 is a stimulator of INS (28, 29). It also associates with proteins regulating food intake, such as pro-opiomelanocortin (POMC) and ghrelin (GHRL) (28–31) (**Figure 6B**). UBS3B associated with EGFR (part of the epidermal growth factor family), CBL (critical for cell signalling pathways), and UBC (which helps regulate cell stress) (**Figure 6C**). ACADV had the largest network of associations, including many proteins associated with fatty acid metabolism (ACOX1, CPT2, and other proteins within the thiolase family (enzymes which break down fatty acids) ACAA1 and ACAA2) (**Figure 6A**). However, Galectin_4 and PRS57 had no protein networks which survived our criteria (high confidence, and from sources detailed in **Figure 6** footnote).

**Figure 6.**
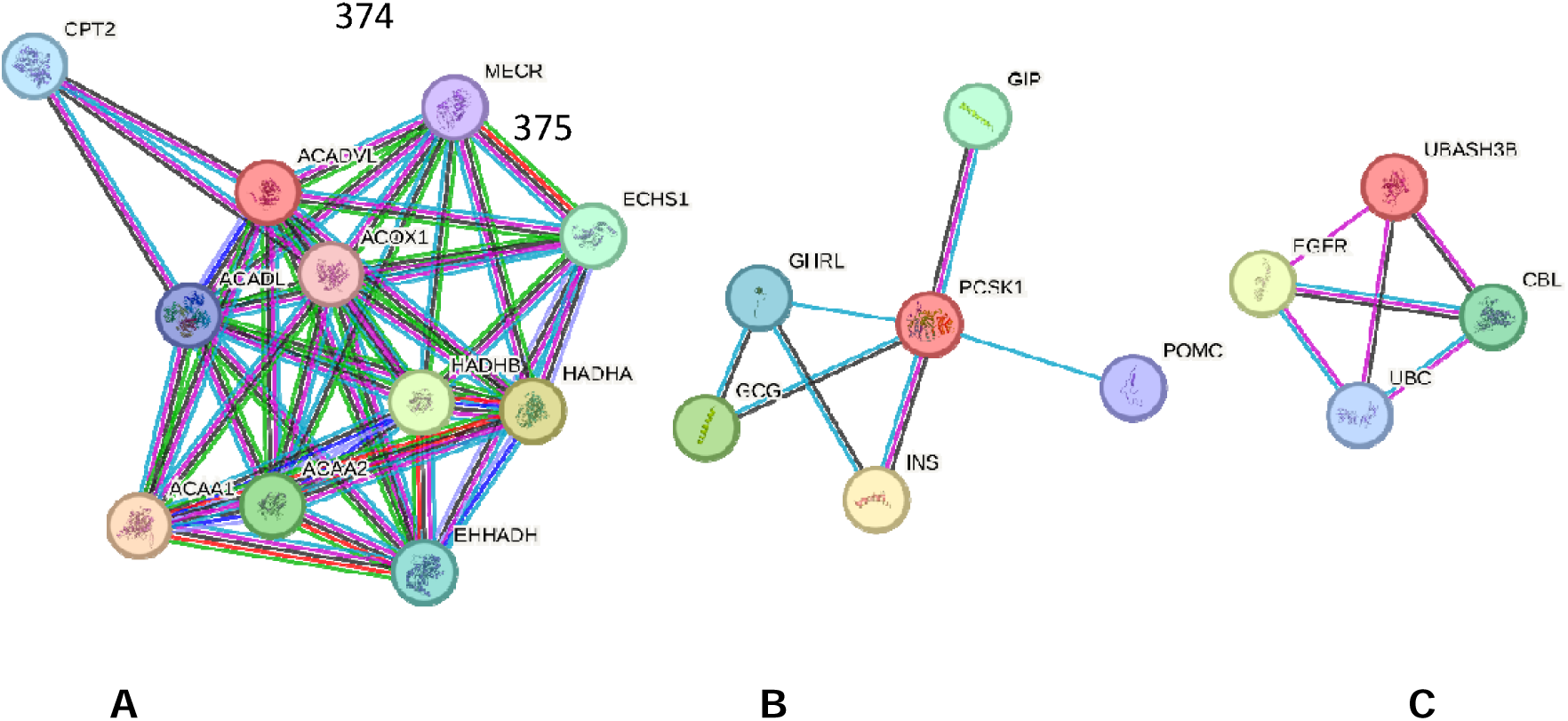
Protein-protein associations - proteins with evidence from both discovery, replication, and colocazation analyses. Different line colours represent different sources of evidence for these predictions – known associations from curated databases (light blue) and experimentally determined (pink); predicted associations from gene neighbourhood (green), gene fusions (red), gene co-occurrence (dark blue), co-expression, and protein homology (lilac). Associations are for proteins jointly contributing to a shared function, rather than specifically binding to each other. Splice isoforms or post-translational modifications are collapsed, i.e. each node represents all the proteins produced by a single, protein-coding gene locus. The physical distances between two nodes along an edge in a graph have no meaning.

## Discussion

We have used two-sample MR to explore the causal effect of maternal pregnancy and fetal plasma proteins on offspring birth weight. We found evidence of potential causal effects, unlikely to be explained by chance or confounding by LD, for 2 maternal circulating proteins (NEC1, Galectin_4) and 3 fetal circulating proteins (NEC1, ACADV, and UBS3B). For the remainder of the discussion, we focus mainly on these proteins which had the strongest evidence across all analyses.

### Proteins with opposing maternal and fetal effects on birthweight

Two proteins, NEC1 and Galectin_4, instrumented by cis-pQTLs, appear to be acting in opposite directions when comparing maternal and fetal MR results. They suggest that more NEC1 from the mother leads to higher birth weight, yet more NEC1 from the fetus leads to lower birth weight – as in the case with insulin and other regulators of fetal growth. This is consistent with previous work showing the role of NEC1 in regulating proinsulin processing (32). Indeed, a new drug (Setmelanotide) being tested to treat obesity is known to perturb PCSK1, because PCSK1 mutations lead to deficient signalling in melanocortin pathways (e.g. because of a lack of α-MSH). Setmelanotide activates MC4R to restore appetite control, addressing genetic obesity caused by disrupted PCSK1 function (33). This is further evidence that our findings could represent molecular targets for birth weight. The STRING protein-protein associations in this study show this link between NEC1 and insulin (INS) and glucagon (GCG) (**Figure 6B**), demonstrating this known function in glucose homeostasis. It is important to note that whilst these protein-protein associations show co-expression (which may be downstream of a certain mechanism), they do not always imply interaction. Furthermore, the different sources of evidence are important when interpreting these interaction networks. We did not include text-mining and gene neighborhood interactions in the STRING networks so we could follow the most robust lines of evidence, and set a high confidence threshold. This may explain why we did not identify any associations for some proteins that replicated and were supported by colocalization, or because proteins like Galectin_4 are poorly characterized (27).

In our analysis, Galectin_4 was shown to the opposite direction of effect to NEC1 on birth weight. More Galectin_4 from the mother leads to lower birthweight and more Galectin_4 from the fetus leads to higher birthweight. Galectin_4 is part of the Galectin family which binds carbohydrates, lactose and sugars (34). Previous work has shown that the Galectin family of β-galactoside binding proteins are important in modulating diverse developmental processes. They contribute to placentation and regulate maternal immune tolerance (34) and have been previously suggested as biomarkers for adverse pregnancy outcomes like gestational diabetes (35) and infertility (34). Other expression studies have shown the importance of Galectins in pregnancy, and the importance of a maternal-fetal balancing act for ideal pregnancy outcomes. For example, lower levels of fetal expression of Galectins 3, 8 and 9 have previously been associated with intrauterine growth restriction (34). However, most of the previous studies of plasma Galectin_4 concentrations in pregnancy have come from mouse or in vitro models (35). It has not been well characterised either inside or outside of pregnancy, demonstrated by the lack of reported associations from the protein-protein association models. Our study provides an additional line of evidence of a potential role of Galectin_4 in fetal growth however there was a lack of colocalization in the fetal effects analysis (34).

PRS57 had strong evidence in the discovery and colocalization analyses for increasing birth weight, however it was the only protein in the maternal analyses which did not replicate. Whilst other members of the serine protease family (a large family of protein-cleaving enzymes) appear critical in pregnancy (e.g. high levels of other serine protease proteins are associated with preeclampsia (36)), to our knowledge, PRS57 has not been previously linked with birth weight, or other disorders of pregnancy. In contrast, PTN9 and ULK3 had evidence from discovery and replication, but not from colocalization. Unfortunately, we had limited power to explore further bias in MR results due to confounding by LD, but high H_3_ results suggest this was down to the traits not sharing the same causal variant, although it could also be related to violations of coloc’s assumptions due to multiple causal variants.

### Proteins with fetal-specific effects on birthweight

UBS3B and ACADV had good evidence from discovery, replication, and colocalization. In our analysis, we found higher UBS3B lowered birth weight. UBS3B is a protein that is not yet well characterised for its role in pregnancy, but known to be associated with cellular processes for mammalian development (37). ACADV (very long-chain acyl-coenzyme A dehydrogenase, also known as VCLAD) catalyses the first step in beta-oxidation of long-chain fatty acids to release energy (38).

In our fetal analysis, we found evidence that higher fetal cis-pQTL instrumented sLeptin_R reduced birth weight. sLeptin_R has a well-established role in energy metabolism, body weight, and is secreted in fetal tissue and the placenta (39). The strong birth weight-associated locus near *CCNL1* from previous GWAS is also associated with leptin levels in adults, providing orthogonal evidence for a key role of this signalling pathway in fetal growth (40). There was good evidence for replication in our study, however the colocalization results fell slightly short of our threshold (H_4_ 0.65, as opposed to 0.70).

### Strengths and limitations

We have integrated the largest-scale proteomics and birthweight genetic association data to shed light on maternal and fetal molecular mechanisms regulating fetal growth. Furthermore, we have used large scale birth weight data from three large European cohorts (EGG, UK Biobank and MoBa) for our discovery, replication analyses and meta analyses. We have undertaken sensitivity analyses, including checking for confounding by LD and attempted to further explain the underlying biology of our molecular findings.

In terms of limitations, our analysis has a limited coverage of the proteome due to the use of GWAS data on plasma proteins. For example, because they are not covered in existing proteomic GWAS, we were unable to assess the effects of two key placental expressed proteins - sFlt-1 and PlGF - , which have biological support for affecting fetal growth, and are now used clinically to predict risk of fetal growth restriction and pre-eclampsia in many countries (41–43). To explore effects of these and potentially other placental proteins we would need large proteomic GWAS of maternal pregnancy and cord-blood proteins. It is also likely that there are additional causal proteins that are not placental-/pregnancy-specific, which might be identified in future, where studies have access to even larger proteomics panels, or using cell- or tissue-specific proteomics.

Another potential limitation is related to epitope effects intrinsic to the proteomic technology. Whilst we aimed to reduce the likelihood of misclassification of protein levels through epitope effects (i.e. differences in antigen recognition, rather than protein levels) we cannot rule out some bias from these effects (44). We removed trans-pQTL and only retained cis-pQTL due to the higher possibility of bias due to pleiotropic mechanisms (45). For each protein we only had one SNP, so methods that have been developed to explore bias due to horizontal pleiotropy are not possible.

In using population level proteomics data, a further limitation is that in the exposure data, whilst we can assume in the maternal analysis that the offspring proteins won’t affect the levels of the maternal proteins, in the offspring analysis, the assumption that maternal proteins won’t affect offspring proteins may not hold. However, there is no way to directly test this.

Furthermore, we have assumed that genetic variants identified in protein GWAS conducted in males and non-pregnant women are similarly associated with circulating proteins in fetuses and pregnant women. Ideally, we would test this in studies that have GWAS data and circulating protein levels in women during pregnancy and cord-blood. However, such data is not currently available or only available in small numbers (for example, there is protein data profiled during pregnancy in the Born in Bradford (BiB) cohort, however the Olink coverage is fewer than 453 proteins in 4000 women, with none of these proteins overlapping with our top hits). However we found evidence that some of the proteins (i.e. NEC1, Galectin_4) that our MR results suggest affect birth weight, are expressed in fetal tissue, which provides some support for their relevance to pregnancy and fetal intrauterine mechanisms. Additionally, supporting evidence from other studies, where genetic variants are instrumental variables for social, behavioural, and molecular exposures during pregnancy, reveals consistent associations with those identified in GWAS of both women and men. This suggests that the assumption is unlikely to be violated. Significantly, with respect to this paper, we have previously demonstrated that among 89 associations involving genetic instruments for one or more of 9 amino acids, there was substantial consistency between those GWAS results derived in men and non-pregnant women, to outcomes in women only (46). This consistency was also observed in women who were pregnant at the time of amino acid measurement for 67 out of the 89 associations (75%). In the remaining 22 associations where heterogeneity was evident, it was likely attributed to the poor quality of genetic imputation for certain variants in the pregnancy study (46).

### Conclusions

We found strong evidence for causal effects of several proteins on offspring birth weight across a range of analyses, highlighting proteins with opposing maternal and offspring effects (NEC1, Galectin_4), and fetal (sLeptin_R, UBS3B, ACADV) effects. These proteins are involved in multiple biological processes governing glucose homeostasis, energy metabolism, endothelial function and adipocyte differentiation. These findings provide new insights into maternal and fetal molecular mechanisms regulating fetal growth.

## Abbreviations

EGG: Early Growth Genetics consortium
eQTL: expression quantitative trait loci
GWAS: genome-wide association study
LD: linkage disequilibrium
MoBa: Norwegian Mother, Father and Child Cohort Study
MR: Mendelian Randomisation
PP: predicted probability
pQTL: protein quantitative trait loci
SNP: single nucleotide polymorphism
UKBB: UK Biobank

## Declarations

### Ethics approval and consent to participate

All human research was approved by the relevant institutional review boards and conducted according to the Declaration of Helsinki. Participants of all studies in the Early Growth Genetics (EGG) consortium provided written informed consent. The UK Biobank has approval from the North West Multi-Centre Research Ethics Committee, which covers the United Kingdom. The establishment of the Norwegian Mother Father and Child (MoBa) birth cohort and initial data collection was based on a license from the Norwegian Data Protection Agency and approval from The Regional Committees for Medical and Health Research Ethics. The MoBa cohort is currently regulated by the Norwegian Health Registry Act. The current study was approved by The Regional Committees for Medical and Health Research Ethics (223273).

### Consent for publication

NA

## Availability of data and materials

### Data availability

Data on birth weight in our discovery analysis was contributed by the EGG Consortium using the UK Biobank Resource and was downloaded from www.egg-consortium.org. The genotype and phenotype data are available on application from the UK Biobank (http://www.ukbiobank.ac.uk/). Individual cohorts participating in the EGG Consortium should be contacted directly as each cohort has different data access policies. The exposure summary data from which genetic instruments were selected for pQTL are available from EpiGraphDB pQTL browser (https://epigraphdb.org/pqtl). Data from MoBa are available from the Norwegian Institute of Public Health after application to the MoBa Scientific Management Group (see its website https://www.fhi.no/en/op/data-access-from-health-registries-health-studies-and-biobanks/data-access/applying-for-access-to-data/ for details).

## Acknowledgments

We gratefully acknowledge the participants in the studies, who were directly or indirectly participating, via consortia summary data, contributed to this research. We also are grateful to researchers who made their data open access. The Norwegian Mother, Father and Child Cohort Study is supported by the Norwegian Ministry of Health and Care Services and the Ministry of Education and Research. We are grateful to all the participating families in Norway who take part in this on-going cohort study. We thank the Norwegian Institute of Public Health (NIPH) for generating high-quality genomic data. This research is part of the HARVEST collaboration, supported by the Research Council of Norway (#229624). We also thank the NORMENT Centre for providing genotype data, funded by the Research Council of Norway (#223273), South East Norway Health Authorities and Stiftelsen Kristian Gerhard Jebsen. We further thank the Center for Diabetes Research, the University of Bergen for providing genotype data and performing quality control and imputation of the data funded by the ERC AdG project SELECTionPREDISPOSED, Stiftelsen Kristian Gerhard Jebsen, Trond Mohn Foundation, the Research Council of Norway, the Novo Nordisk Foundation, the University of Bergen, and the Western Norway Health Authorities. This work was carried out using the computational facilities of the Advanced Computing Research Centre, University of Bristol - http://www.bristol.ac.uk/acrc/ and the TSD (Tjeneste for Sensitive Data) facilities, owned by the University of Oslo, operated and developed by the TSD service group at the University of Oslo, IT Department (USIT). The computations on this platform were performed on resources provided by Sigma2 - the National Infrastructure for High-Performance Computing and Data Storage in Norway.

## Funding

This work was supported by the University of Bristol and UK Medical Research Council ( MC_UU_00032/05), the US National Institute for Health (R01 DK10324), the European Research Council via Advanced Grant 101021566, the British Heart Foundation (AA/18/7/34219 and CS/16/4/32482) and the National Institute of Health Research Bristol Biomedical Research Centre at University Hospitals Bristol NHS Foundation Trust and the University of Bristol.

RMF and RNB were funded by a Wellcome Trust and Royal Society Sir Henry Dale Fellowship (WT104150). RMF is supported by a Wellcome Senior Research Fellowship (WT220390). This research was supported by the National Institute for Health and Care Research Exeter Biomedical Research Centre. This research was funded in part by the Wellcome Trust [WT220390]. J.Z. is supported by grants from the National Key Research and Development Program of China (2022YFC2505203). MCB is funded by a University of Bristol Vice Chancellor’s Fellowship. EC is supported by funding from the Research Council of Norway (#274611) and the South-Eastern Norway Regional Health Authority (#2021045). The views expressed in this paper are those of the authors and not necessarily those of the any of the funders listed above.

For open access, the authors have applied a CC BY public copyright licence to any Author Accepted Manuscript version arising from this submission.

## Author contributions

Study conception: DAL, MCB, RMF; Analysis: NM, MCB; Manuscript drafting: NM; Manuscript revisions: NM, MCB, DAL, AFS, MA, RB, RMF, TB, TRG, JZ, DE, DH, GC, EC, MM

## Code availability

All code is available on the GitHub repo https://github.com/MRCIEU/MR-PREG

## Supplementary

**Supplementary figure 1.**
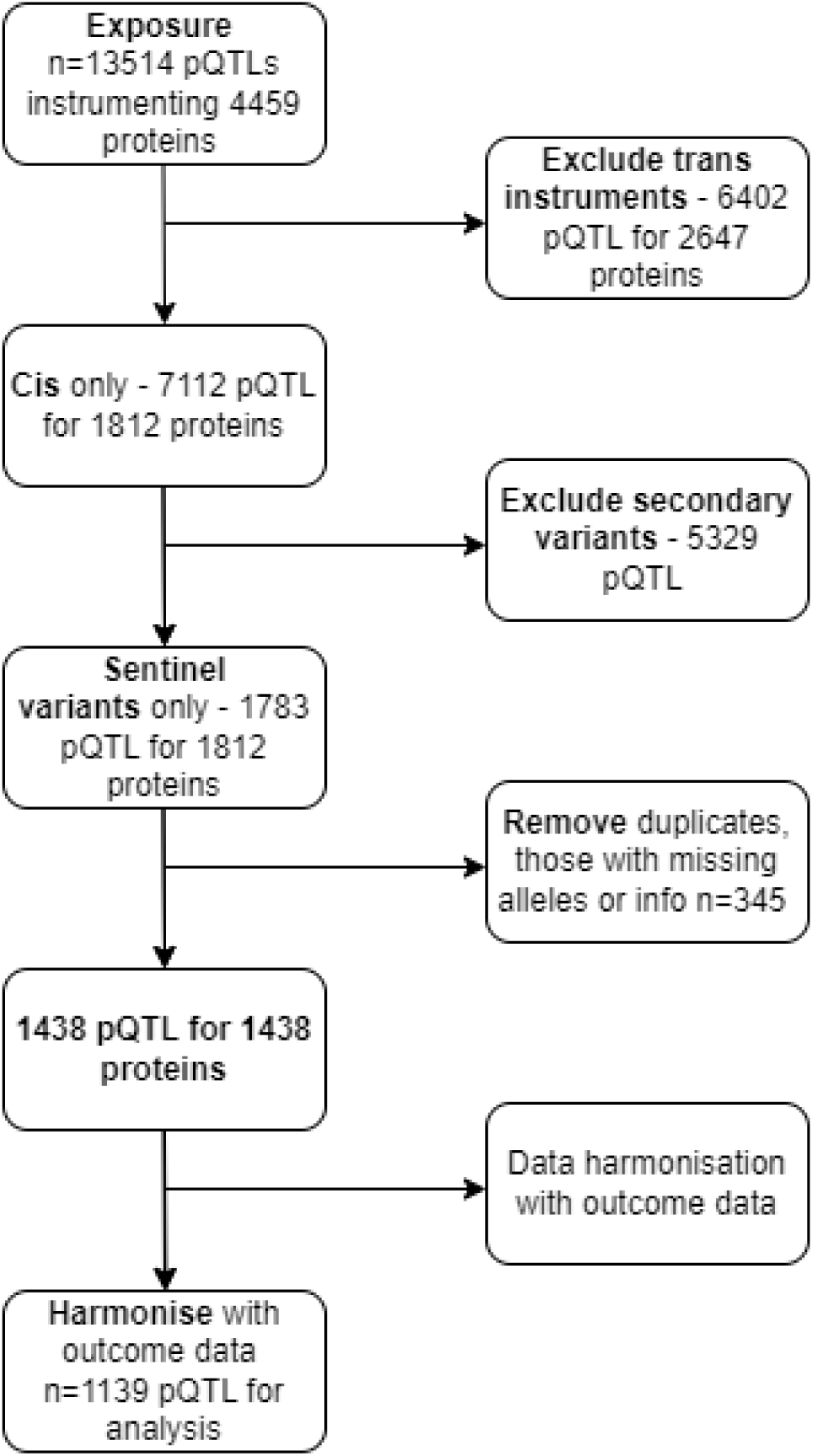
a workflow of instruments (pQTL) used in our analysis. Abbreviations: pQTL, protein quantitative trait loci

**Supplementary figure 2.**
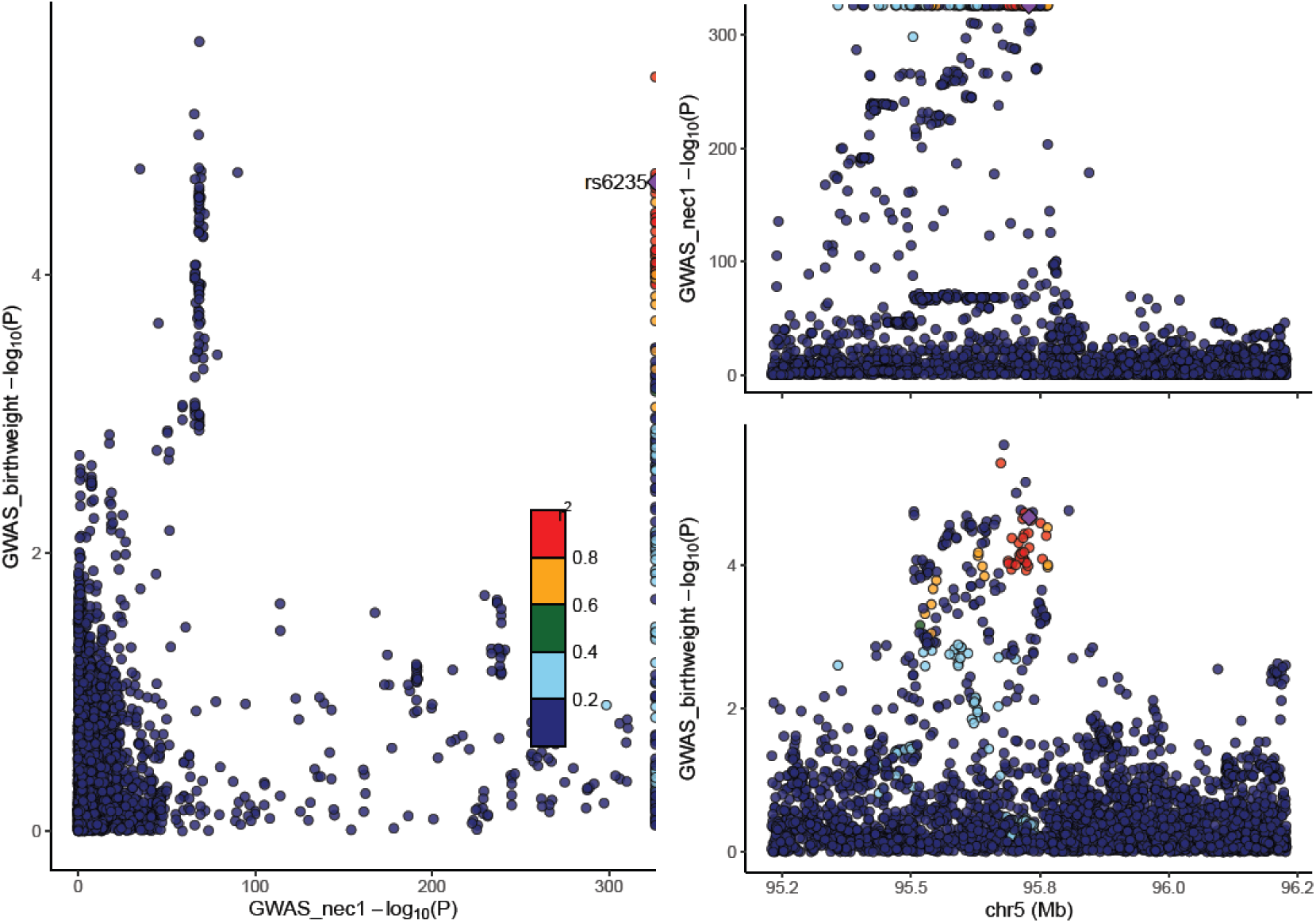
Genetic associations with NEC1 and offspring birth weight in maternal analysis. Each data point represents one genetic variant. The purple diamond represents the selected pQTL. Colours indicate the R^2^ values for strength of linkage disequilibrium between the genetic variant and the pQTL. In the left panel, the plot shows the correlation between log 10 p-values for the genetic association with protein levels (x axis) and birth weight (y axis). The right panels show recombination plots for the protein (top panel) and birth weight (bottom panel) with log10 p-values in the y axis and chromosome positions in the x axis.

**Supplementary figure 3.**
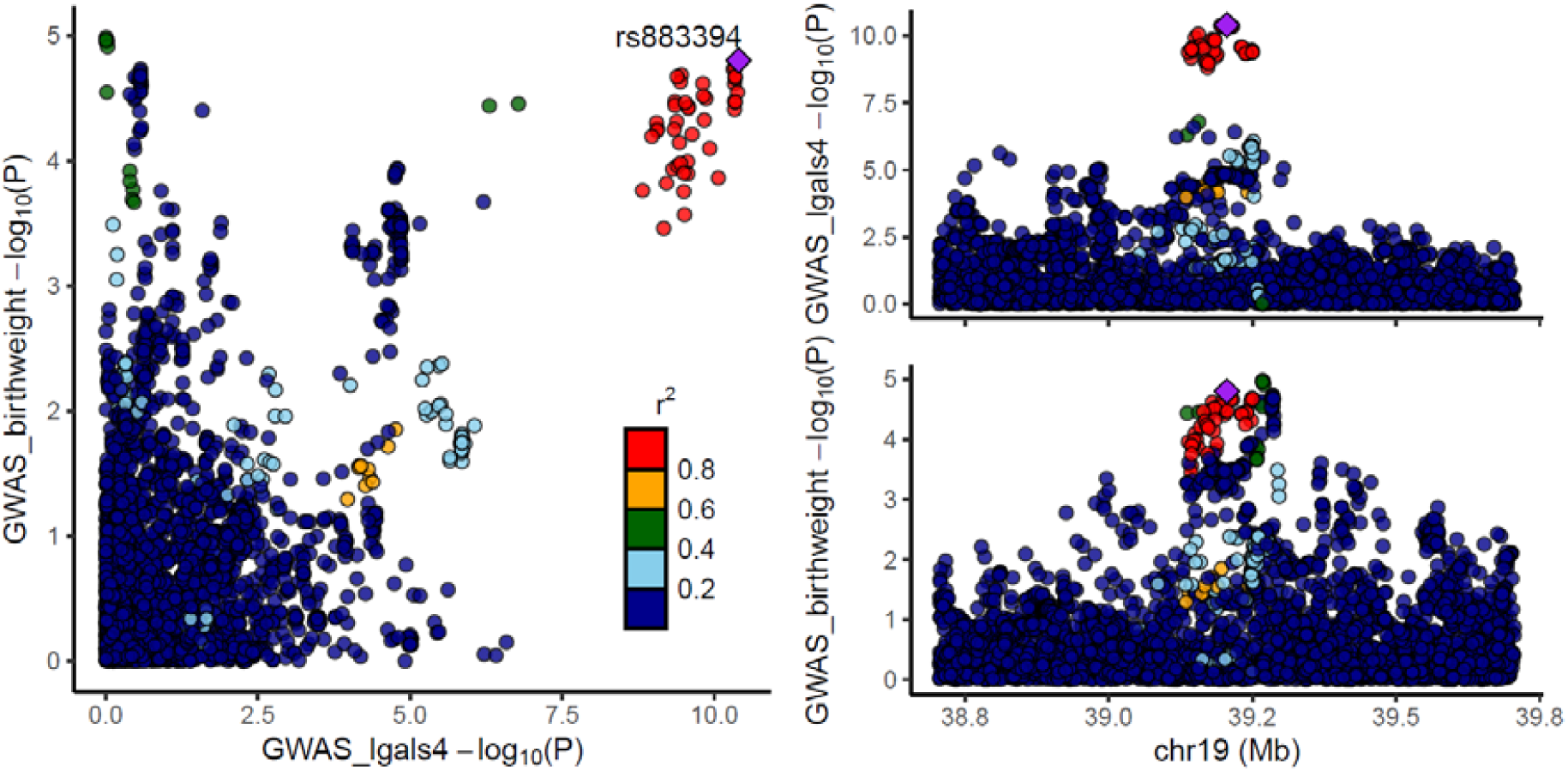
Genetic associations with Galectin_4 and offspring birth weight in maternal analysis. Each data point represents one genetic variant. The purple diamond represents the selected pQTL. Colours indicate the R^2^ values for strength of linkage disequilibrium between the genetic variant and the pQTL. In the left panel, the plot shows the correlation between log 10 p-values for the genetic association with protein levels (x axis) and birth weight (y axis). The right panels show recombination plots for the protein (top panel) and birth weight (bottom panel) with log10 p-values in the y axis and chromosome positions in the x axis.

**Supplementary figure 4.**
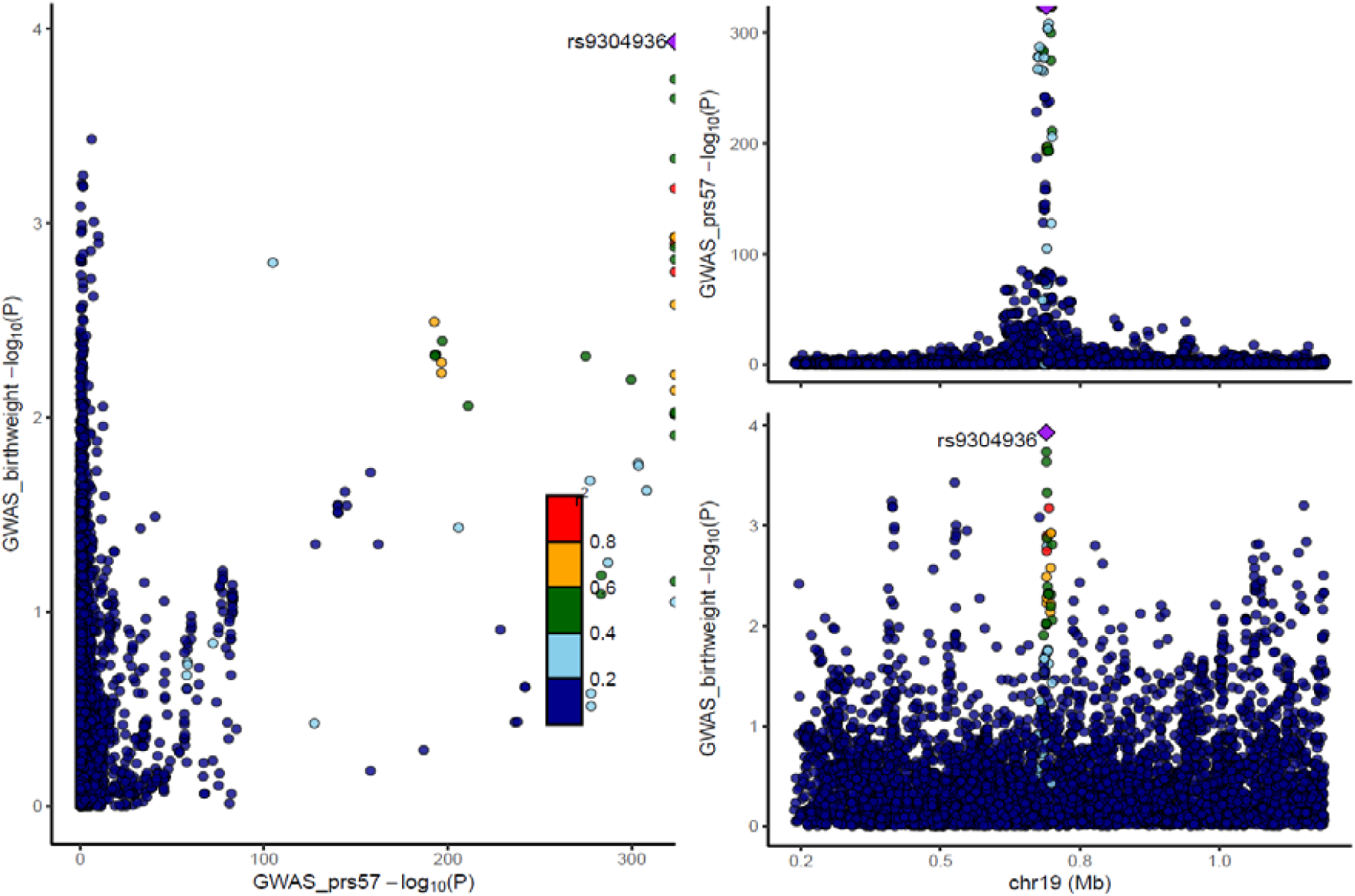
Genetic associations with PRS57 and offspring birth weight in maternal analysis. Each data point represents one genetic variant. The purple diamond represents the selected pQTL. Colours indicate the R^2^ values for strength of linkage disequilibrium between the genetic variant and the pQTL. In the left panel, the plot shows the correlation between log 10 p-values for the genetic association with protein levels (x axis) and birth weight (y axis). The right panels show recombination plots for the protein (top panel) and birth weight (bottom panel) with log10 p-values in the y axis and chromosome positions in the x axis.

**Supplementary figure 5.**
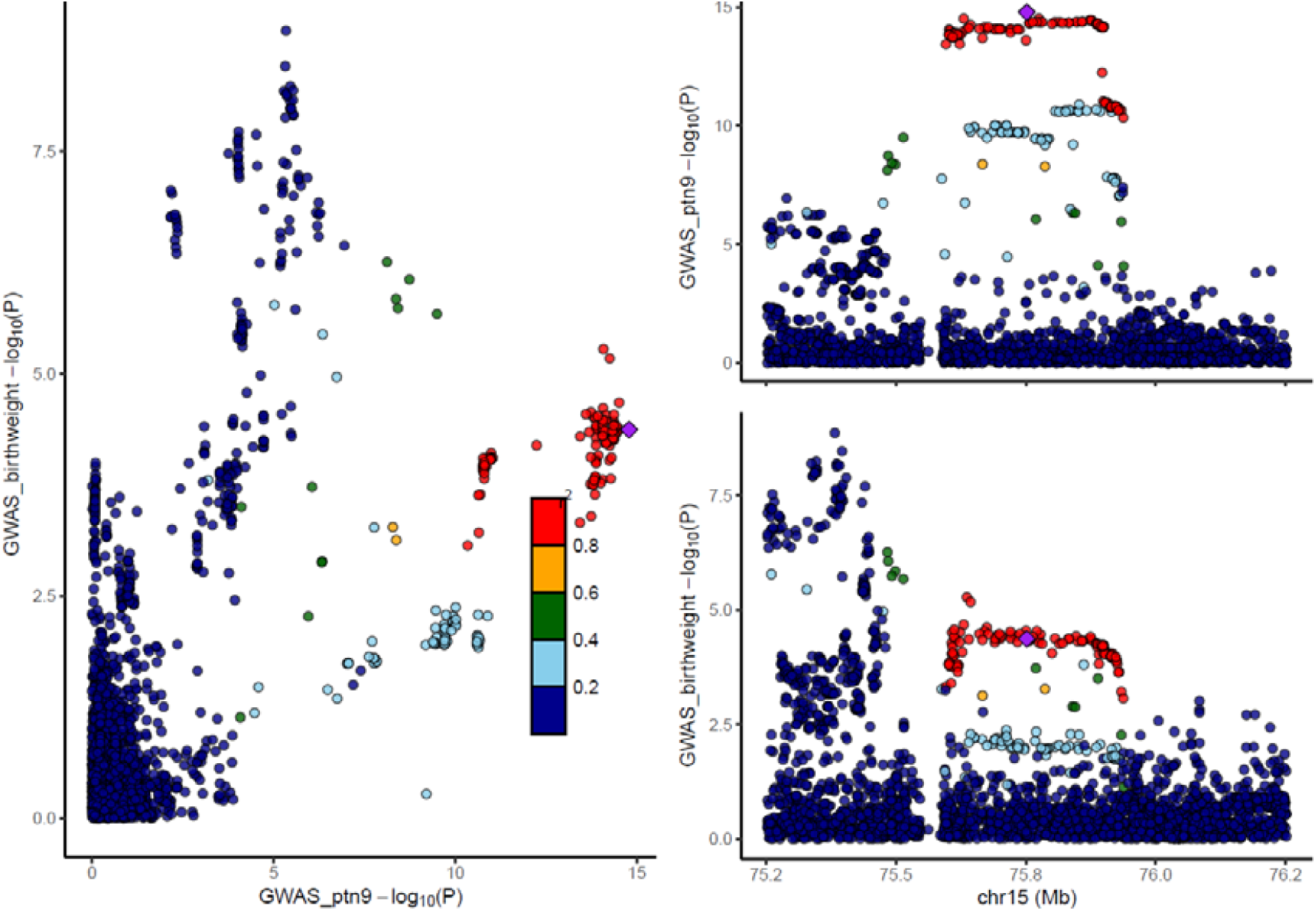
Genetic associations with PTN9 and offspring birth weight in offspring analyses. Each data point represents one genetic variant. The purple diamond represents the selected pQTL. Colours indicate the R^2^ values for strength of linkage disequilibrium between the genetic variant and the pQTL. In the left panel, the plot shows the correlation between log 10 p-values for the genetic association with protein levels (x axis) and birth weight (y axis). The right panels show recombination plots for the protein (top panel) and birth weight (bottom panel) with log10 p-values in the y axis and chromosome positions in the x axis.

**Supplementary figure 6.**
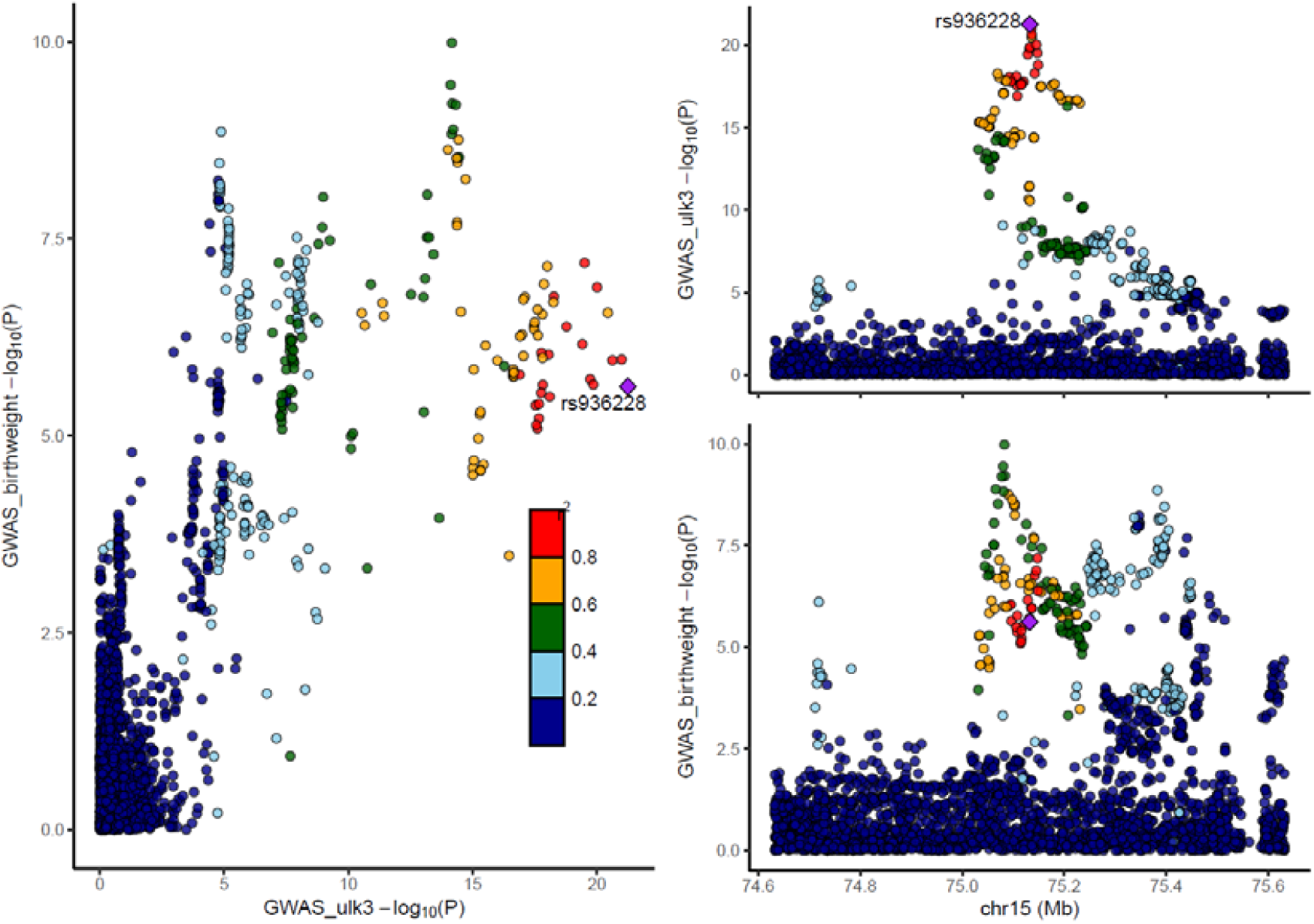
Genetic associations with ULK3 and offspring birth weight in maternal analysis. Each data point represents one genetic variant. The purple diamond represents the selected pQTL. Colours indicate the R^2^ values for strength of linkage disequilibrium between the genetic variant and the pQTL. In the left panel, the plot shows the correlation between log 10 p-values for the genetic association with protein levels (x axis) and birth weight (y axis). The right panels show recombination plots for the protein (top panel) and birth weight (bottom panel) with log10 p-values in the y axis and chromosome positions in the x axis.

**Supplementary figure 7.**
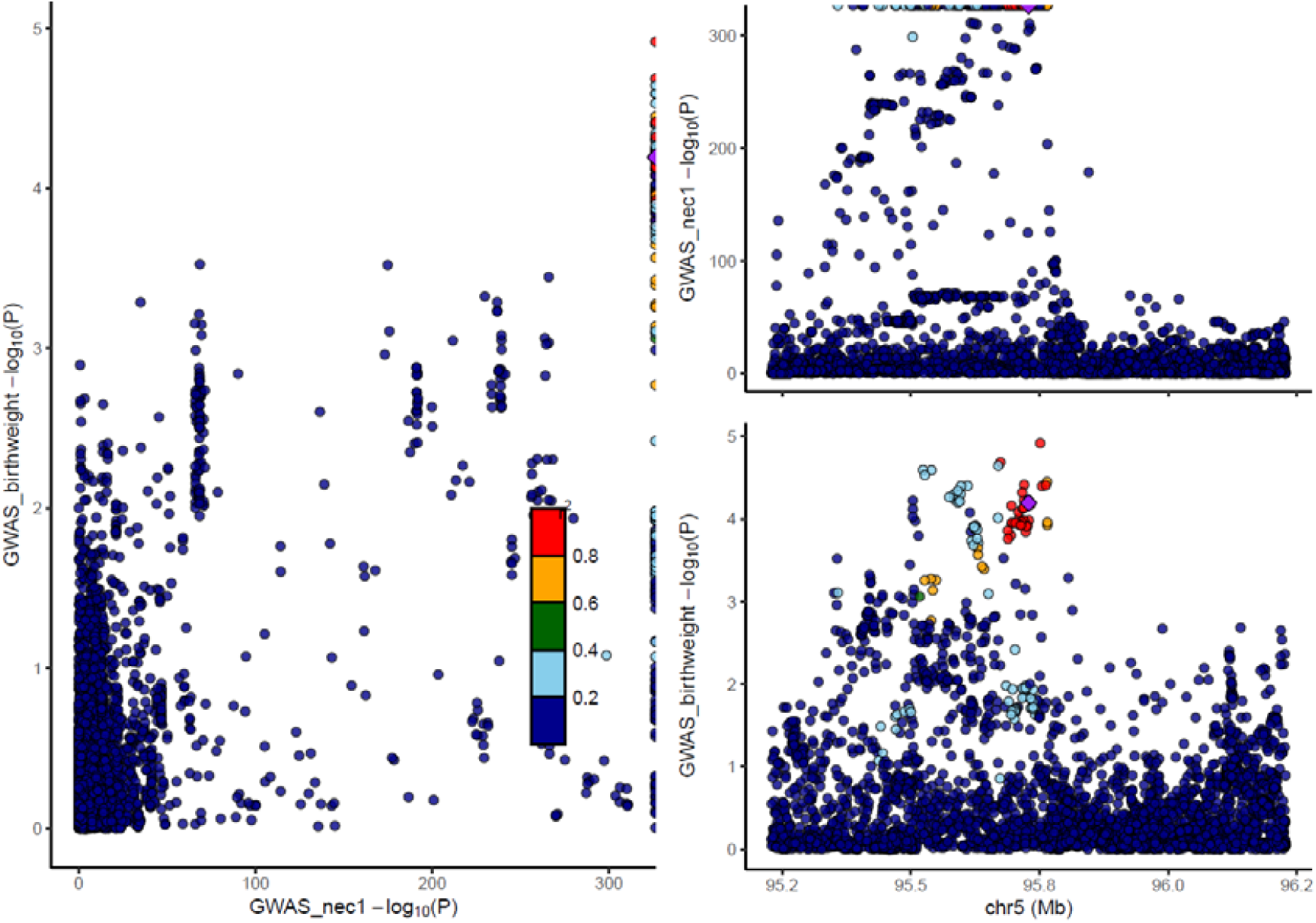
Genetic associations with NEC1 and offspring birth weight in offspring analysis. Each data point represents one genetic variant. The purple diamond represents the selected pQTL. Colours indicate the R^2^ values for strength of linkage disequilibrium between the genetic variant and the pQTL. In the left panel, the plot shows the correlation between log 10 p-values for the genetic association with protein levels (x axis) and birth weight (y axis). The right panels show recombination plots for the protein (top panel) and birth weight (bottom panel) with log10 p-values in the y axis and chromosome positions in the x axis.

**Supplementary figure 8.**
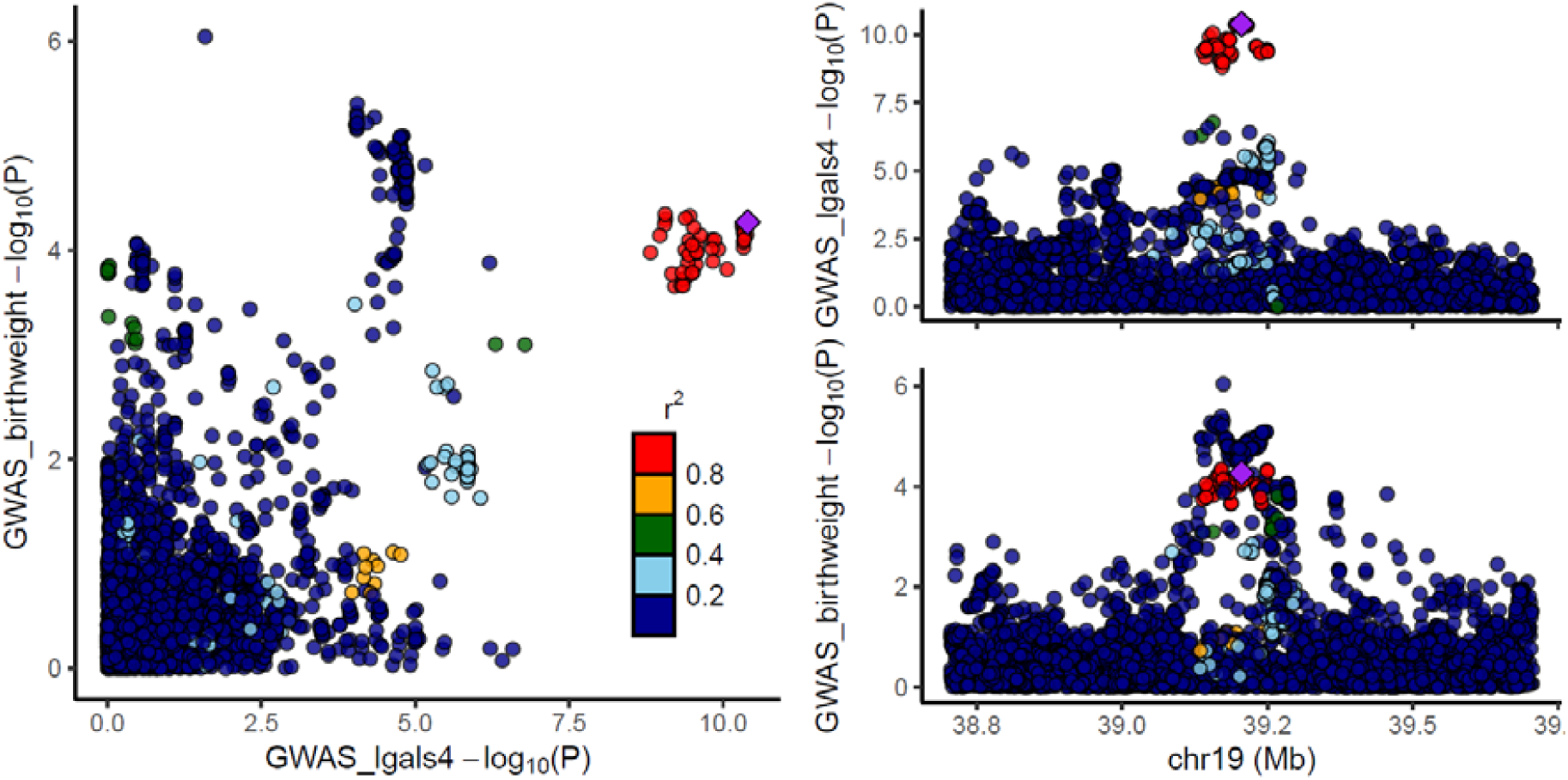
Genetic associations with Galectin_4 and offspring birth weight in offspring analysis. Each data point represents one genetic variant. The purple diamond represents the selected pQTL. Colours indicate the R^2^ values for strength of linkage disequilibrium between the genetic variant and the pQTL. In the left panel, the plot shows the correlation between log 10 p-values for the genetic association with protein levels (x axis) and birth weight (y axis). The right panels show recombination plots for the protein (top panel) and birth weight (bottom panel) with log10 p-values in the y axis and chromosome positions in the x axis.

**Supplementary figure 9.**
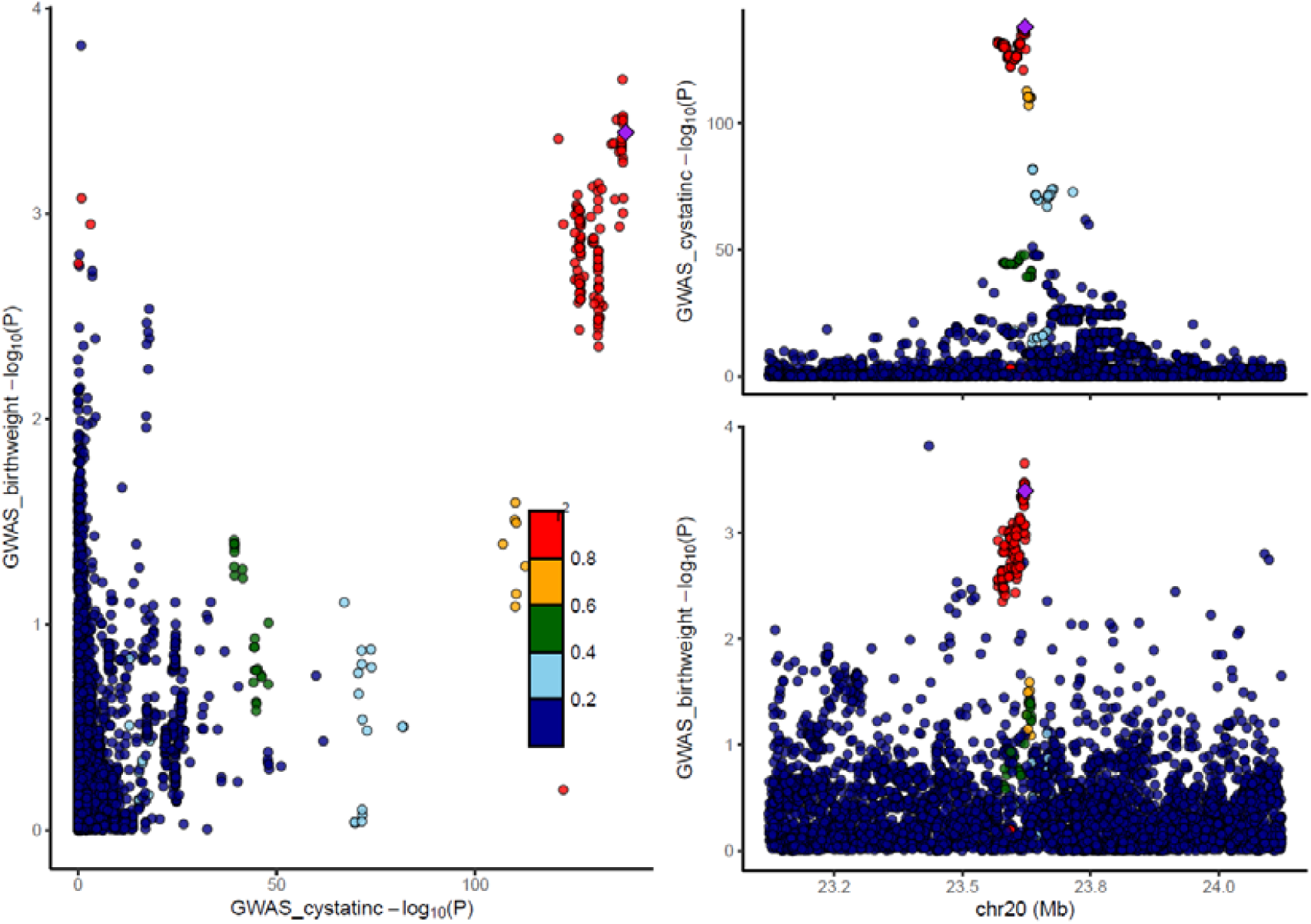
Genetic associations with Cystatin_c and offspring birth weight in offspring analysis. Each data point represents one genetic variant. The purple diamond represents the selected pQTL. Colours indicate the R^2^ values for strength of linkage disequilibrium between the genetic variant and the pQTL. In the left panel, the plot shows the correlation between log 10 p-values for the genetic association with protein levels (x axis) and birth weight (y axis). The right panels show recombination plots for the protein (top panel) and birth weight (bottom panel) with log10 p-values in the y axis and chromosome positions in the x axis.

**Supplementary figure 10.**
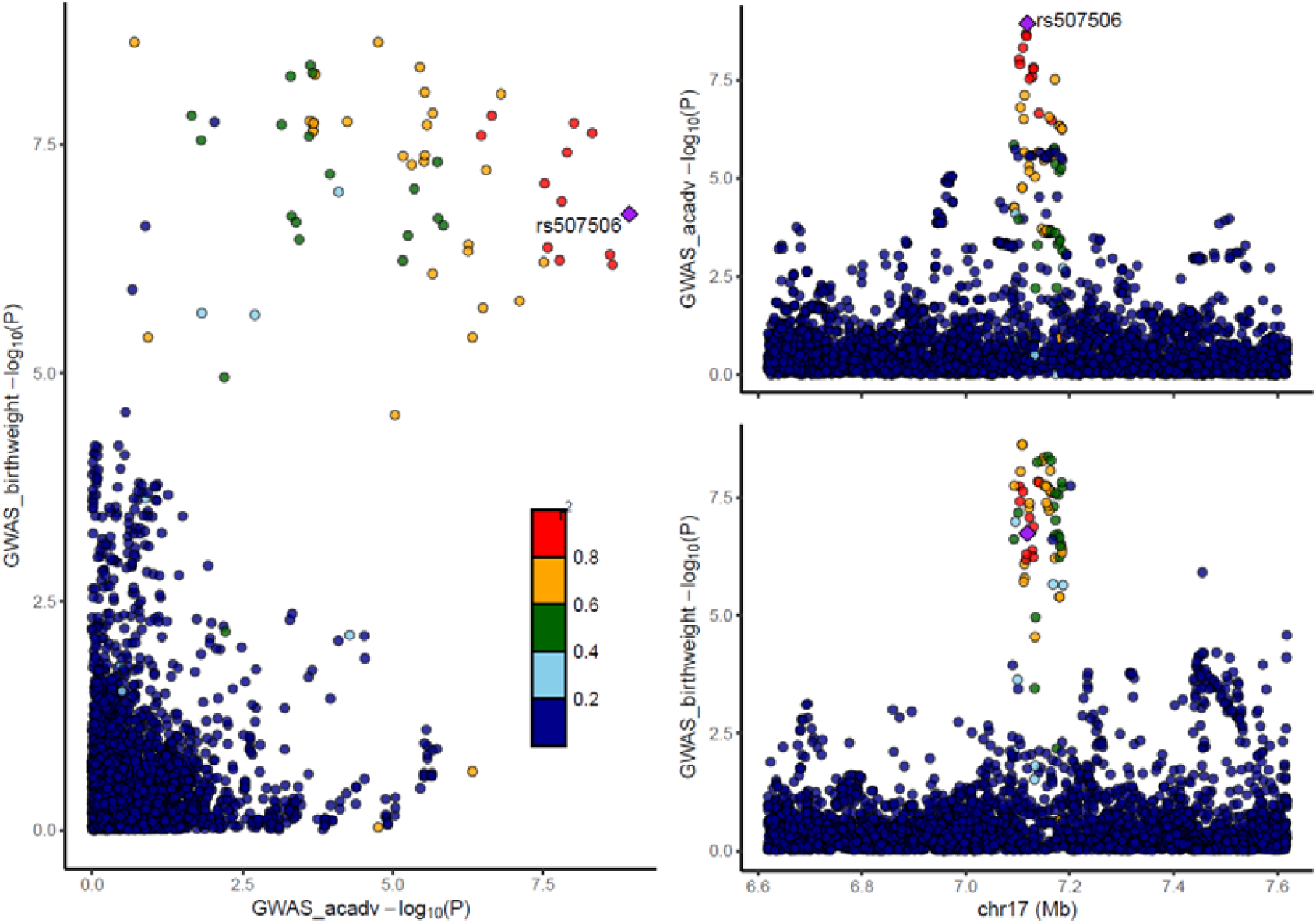
Genetic associations with ACADV and offspring birth weight in offspring analysis. Each data point represents one genetic variant. The purple diamond represents the selected pQTL. Colours indicate the R^2^ values for strength of linkage disequilibrium between the genetic variant and the pQTL. In the left panel, the plot shows the correlation between log 10 p-values for the genetic association with protein levels (x axis) and birth weight (y axis). The right panels show recombination plots for the protein (top panel) and birth weight (bottom panel) with log10 p-values in the y axis and chromosome positions in the x axis.

**Supplementary figure 11.**
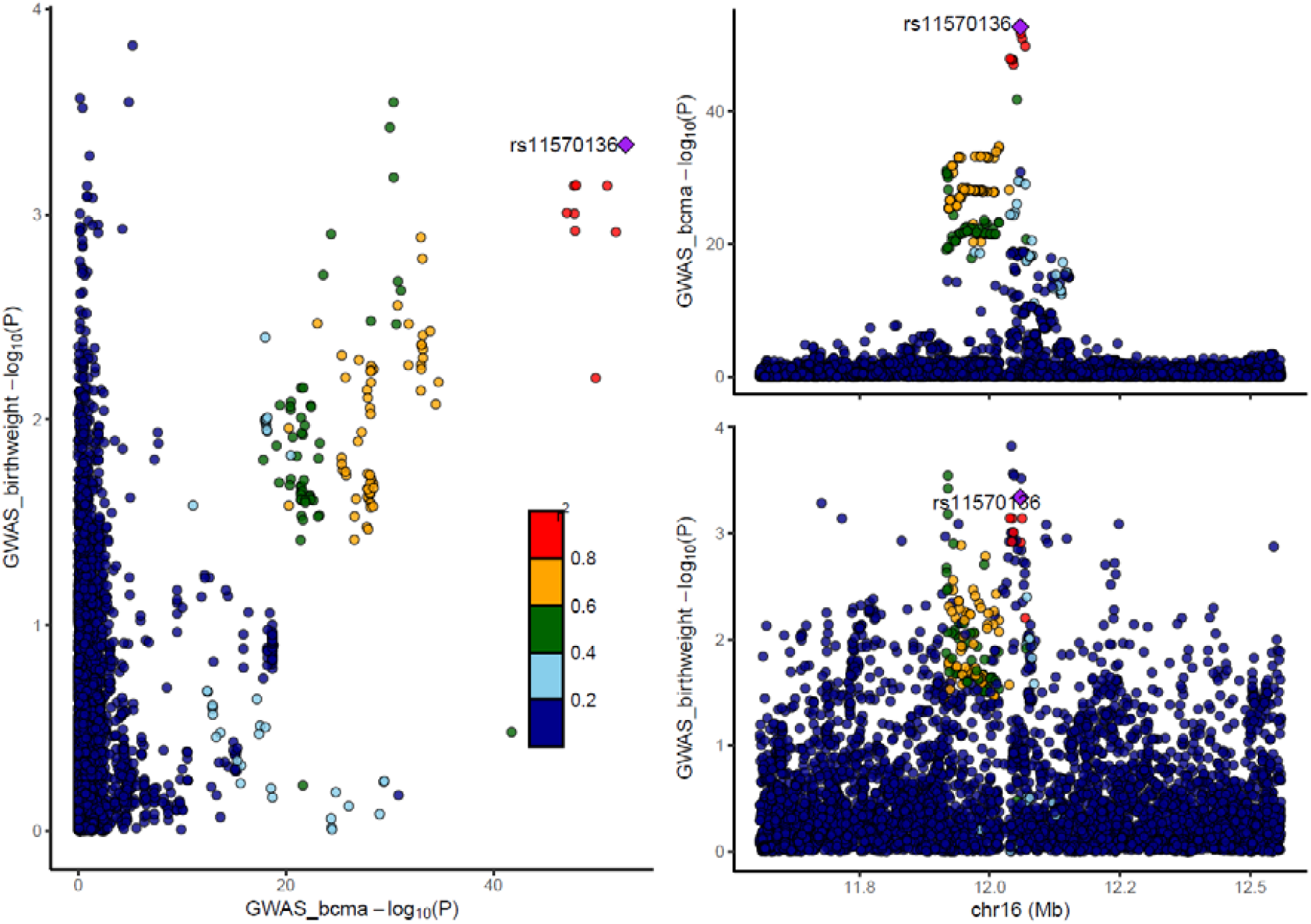
Genetic associations with BCMA and offspring birth weight in offspring analysis. Each data point represents one genetic variant. The purple diamond represents the selected pQTL. Colours indicate the R^2^ values for strength of linkage disequilibrium between the genetic variant and the pQTL. In the left panel, the plot shows the correlation between log 10 p-values for the genetic association with protein levels (x axis) and birth weight (y axis). The right panels show recombination plots for the protein (top panel) and birth weight (bottom panel) with log10 p-values in the y axis and chromosome positions in the x axis.

**Supplementary figure 12.**
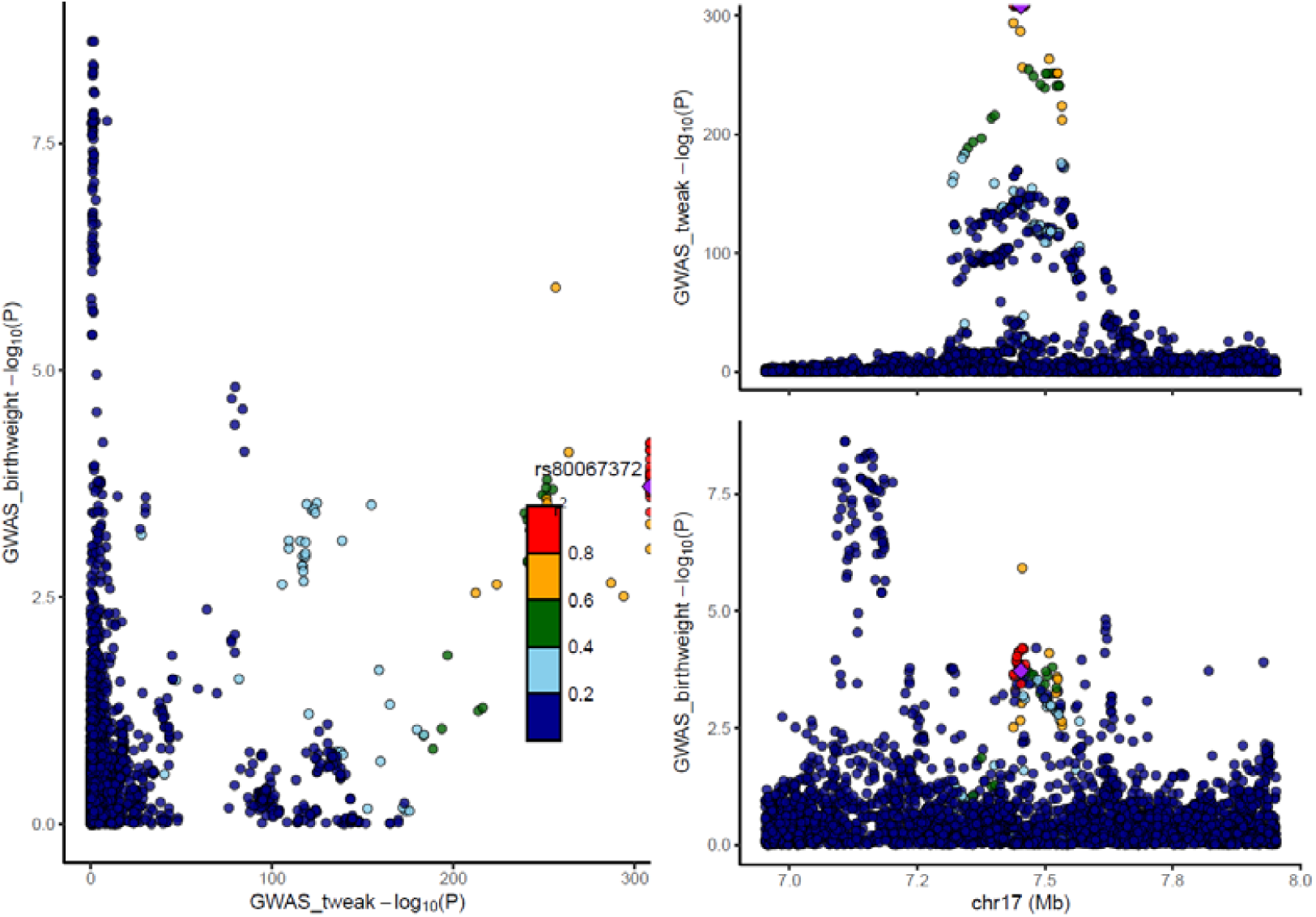
Genetic associations with TWEAK and offspring birth weight in offspring analysis. Each data point represents one genetic variant. The purple diamond represents the selected pQTL. Colours indicate the R^2^ values for strength of linkage disequilibrium between the genetic variant and the pQTL. In the left panel, the plot shows the correlation between log 10 p-values for the genetic association with protein levels (x axis) and birth weight (y axis). The right panels show recombination plots for the protein (top panel) and birth weight (bottom panel) with log10 p-values in the y axis and chromosome positions in the x axis.

**Supplementary figure 13.**
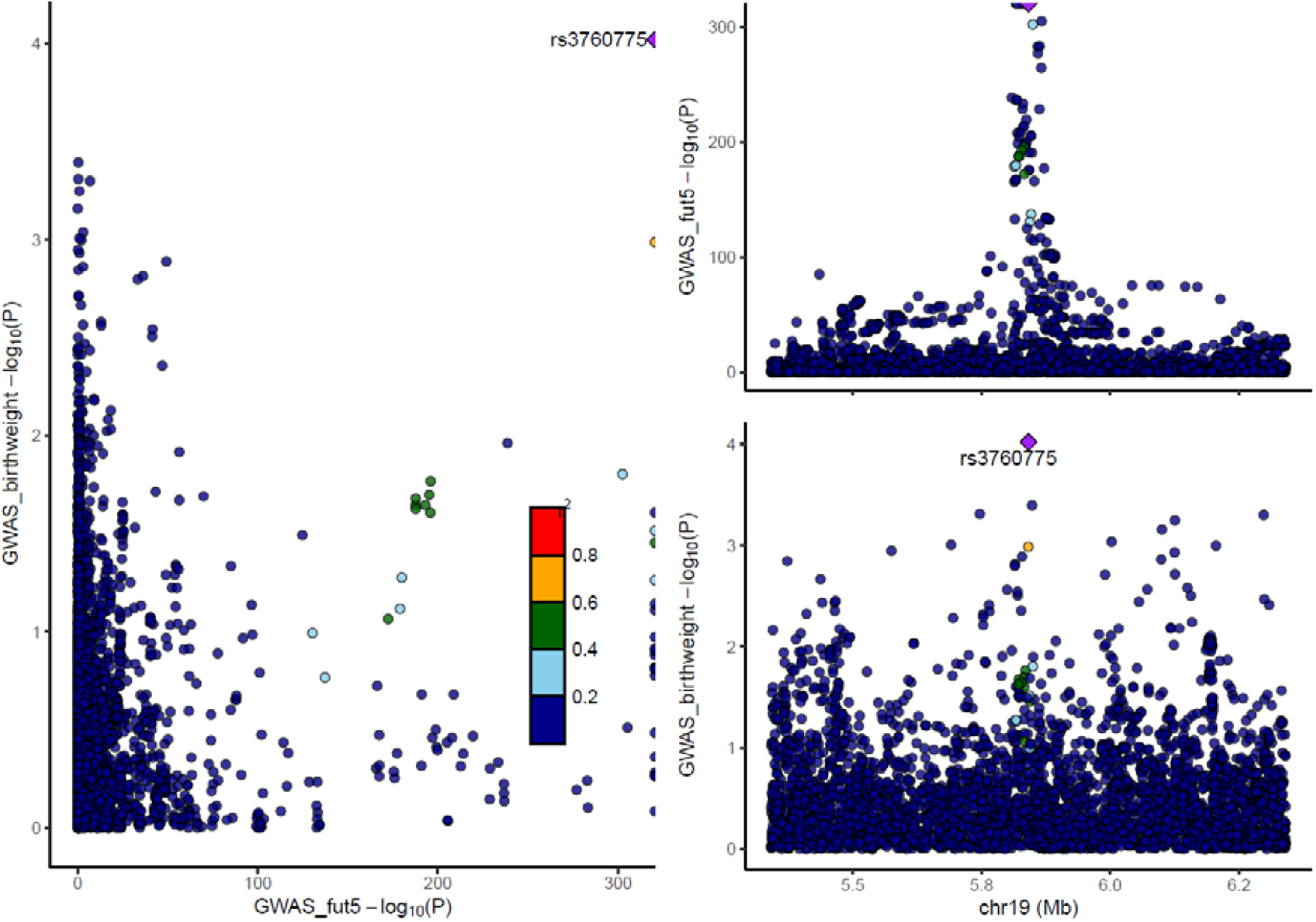
Genetic associations with FUT5 and offspring birth weight in offspring analysis. Each data point represents one genetic variant. The purple diamond represents the selected pQTL. Colours indicate the R^2^ values for strength of linkage disequilibrium between the genetic variant and the pQTL. In the left panel, the plot shows the correlation between log 10 p-values for the genetic association with protein levels (x axis) and birth weight (y axis). The right panels show recombination plots for the protein (top panel) and birth weight (bottom panel) with log10 p-values in the y axis and chromosome positions in the x axis.

**Supplementary figure 14.**
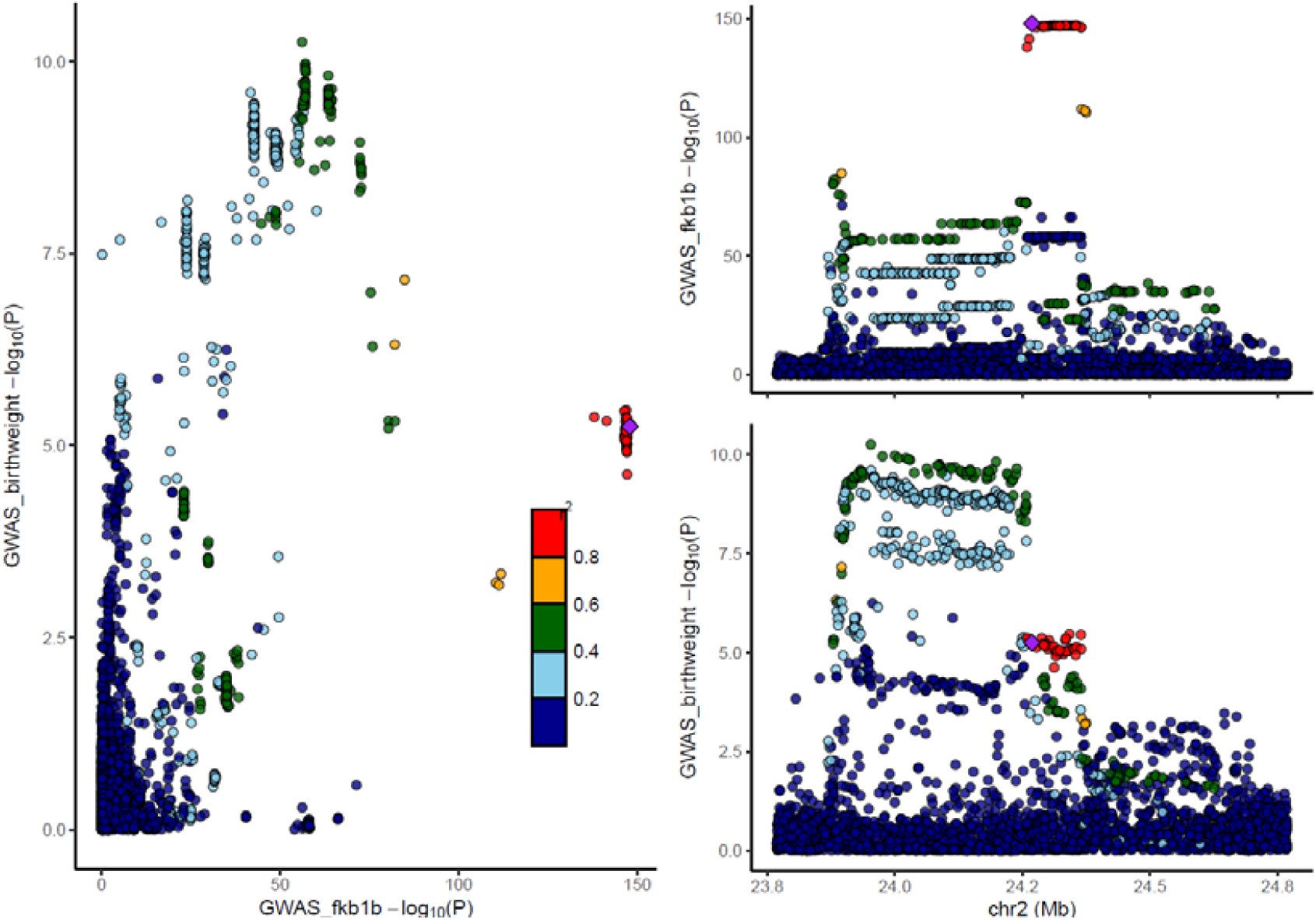
Genetic associations with FKB1B and offspring birth weight in offspring analysis. Each data point represents one genetic variant. The purple diamond represents the selected pQTL. Colours indicate the R^2^ values for strength of linkage disequilibrium between the genetic variant and the pQTL. In the left panel, the plot shows the correlation between log 10 p-values for the genetic association with protein levels (x axis) and birth weight (y axis). The right panels show recombination plots for the protein (top panel) and birth weight (bottom panel) with log10 p-values in the y axis and chromosome positions in the x axis.

**Supplementary figure 15.**
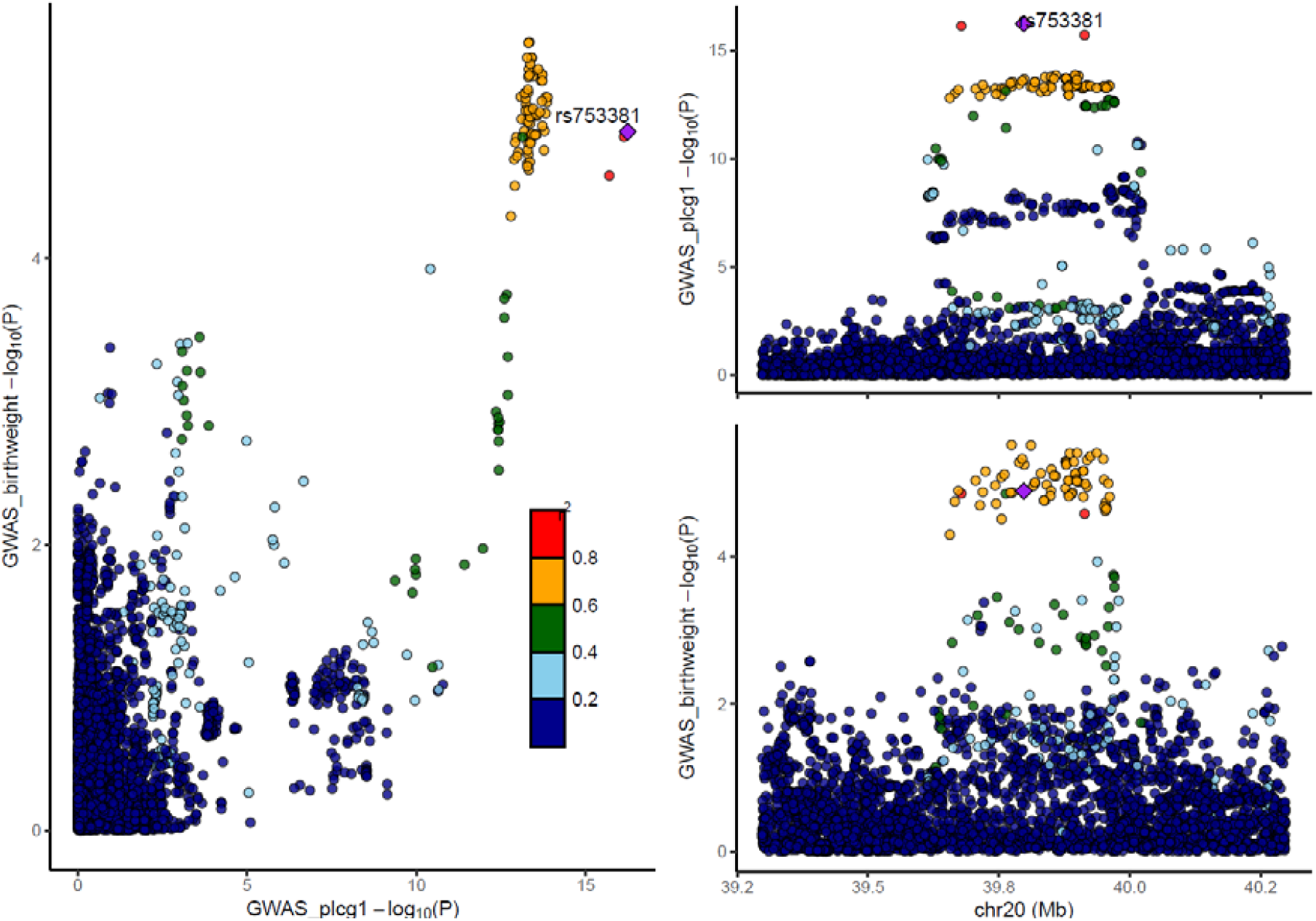
Genetic associations with PLCG1 and offspring birth weight in offspring analysis. Each data point represents one genetic variant. The purple diamond represents the selected pQTL. Colours indicate the R^2^ values for strength of linkage disequilibrium between the genetic variant and the pQTL. In the left panel, the plot shows the correlation between log 10 p-values for the genetic association with protein levels (x axis) and birth weight (y axis). The right panels show recombination plots for the protein (top panel) and birth weight (bottom panel) with log10 p-values in the y axis and chromosome positions in the x axis.

**Supplementary figure 16.**
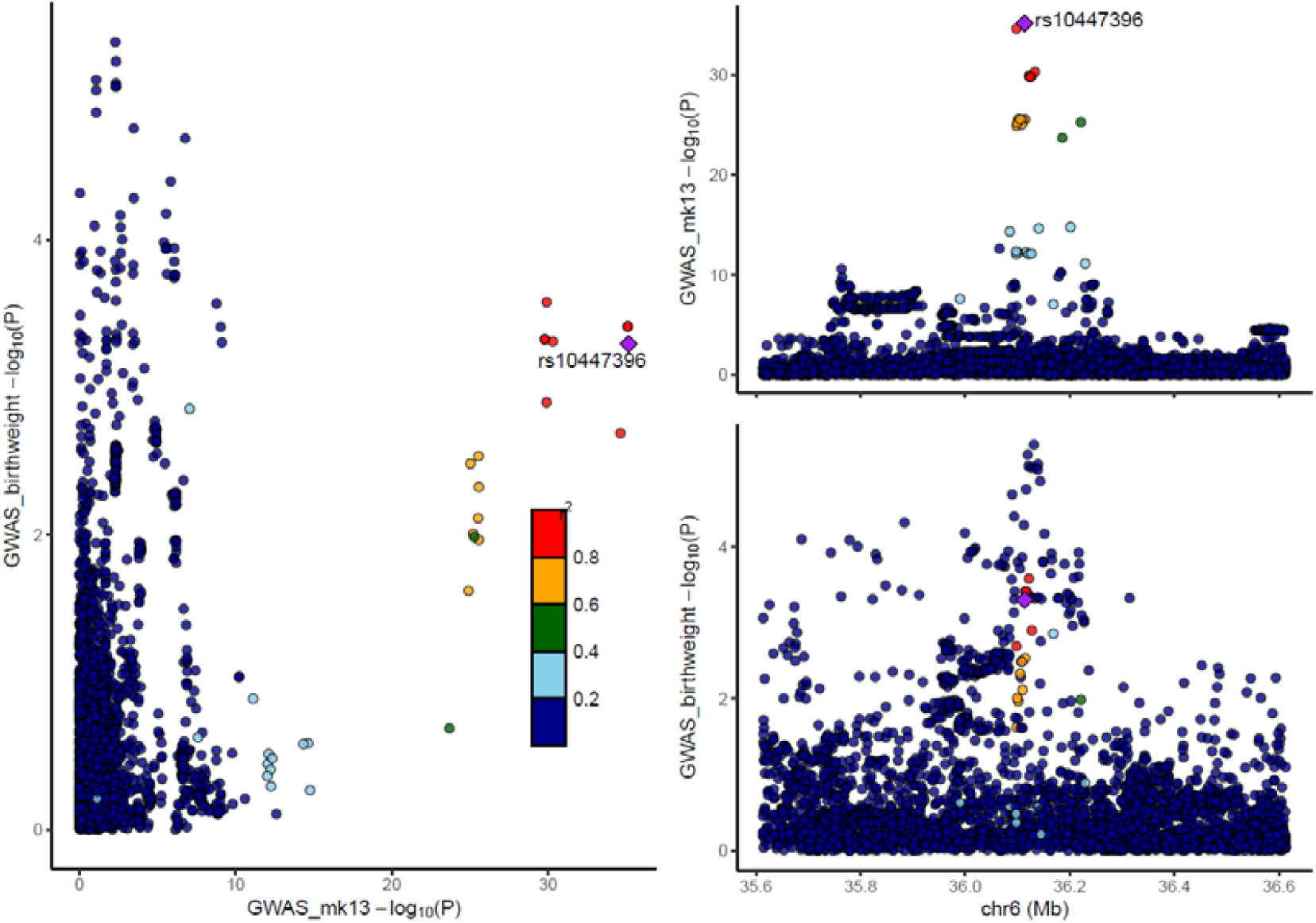
Genetic associations with MK13 and offspring birth weight in offspring analysis. Each data point represents one genetic variant. The purple diamond represents the selected pQTL. Colours indicate the R^2^ values for strength of linkage disequilibrium between the genetic variant and the pQTL. In the left panel, the plot shows the correlation between log 10 p-values for the genetic association with protein levels (x axis) and birth weight (y axis). The right panels show recombination plots for the protein (top panel) and birth weight (bottom panel) with log10 p-values in the y axis and chromosome positions in the x axis.

**Supplementary figure 18.**
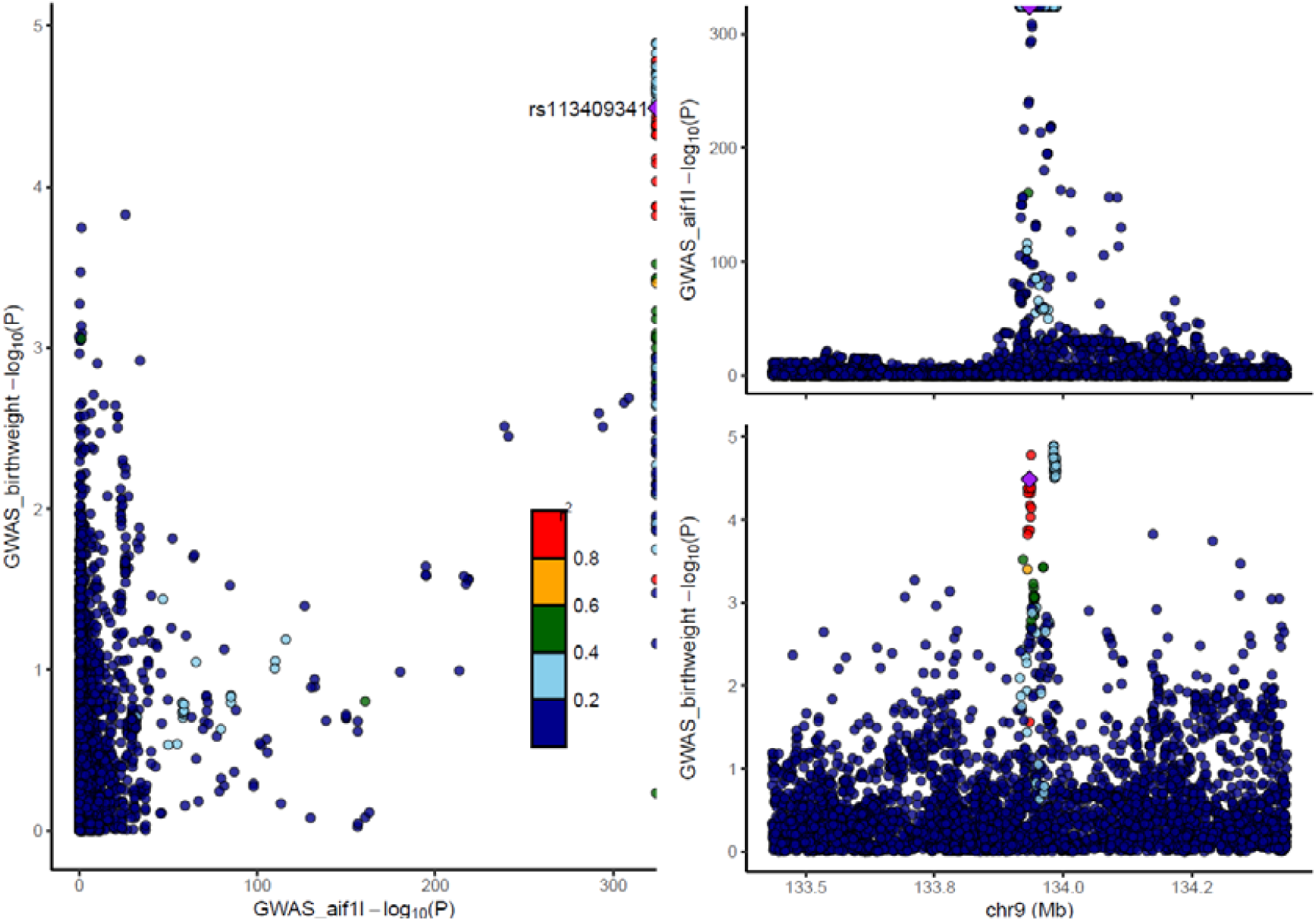
Genetic associations with AIF1L and offspring birth weight in offspring analysis. Each data point represents one genetic variant. The purple diamond represents the selected pQTL. Colours indicate the R^2^ values for strength of linkage disequilibrium between the genetic variant and the pQTL. In the left panel, the plot shows the correlation between log 10 p-values for the genetic association with protein levels (x axis) and birth weight (y axis). The right panels show recombination plots for the protein (top panel) and birth weight (bottom panel) with log10 p-values in the y axis and chromosome positions in the x axis.

**Supplementary figure 19.**
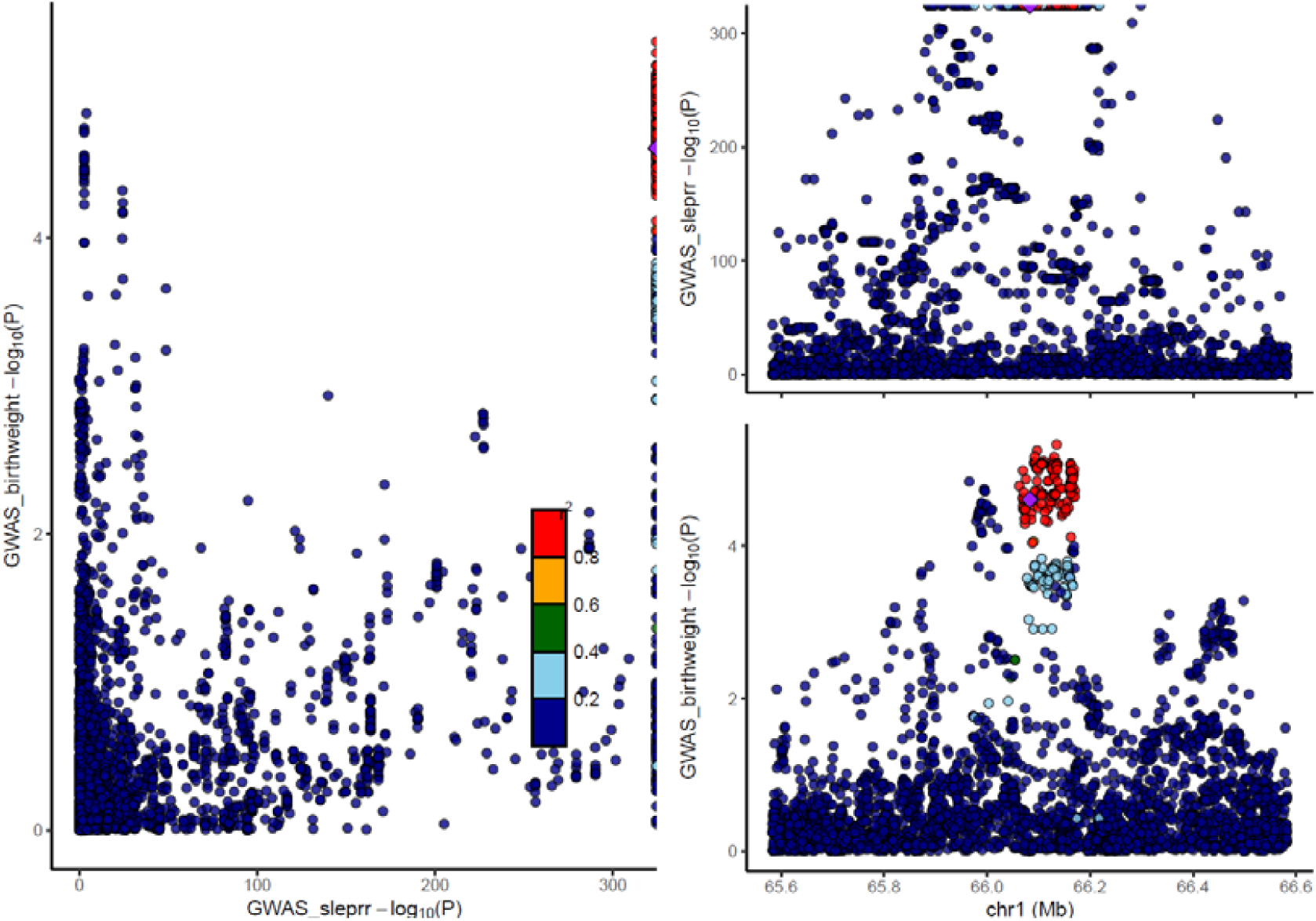
Genetic associations with sLeptin_R and offspring birth weight in offspring analysis. Each data point represents one genetic variant. The purple diamond represents the selected pQTL. Colours indicate the R^2^ values for strength of linkage disequilibrium between the genetic variant and the pQTL. In the left panel, the plot shows the correlation between log 10 p-values for the genetic association with protein levels (x axis) and birth weight (y axis). The right panels show recombination plots for the protein (top panel) and birth weight (bottom panel) with log10 p-values in the y axis and chromosome positions in the x axis.

**Supplementary figure 20.**
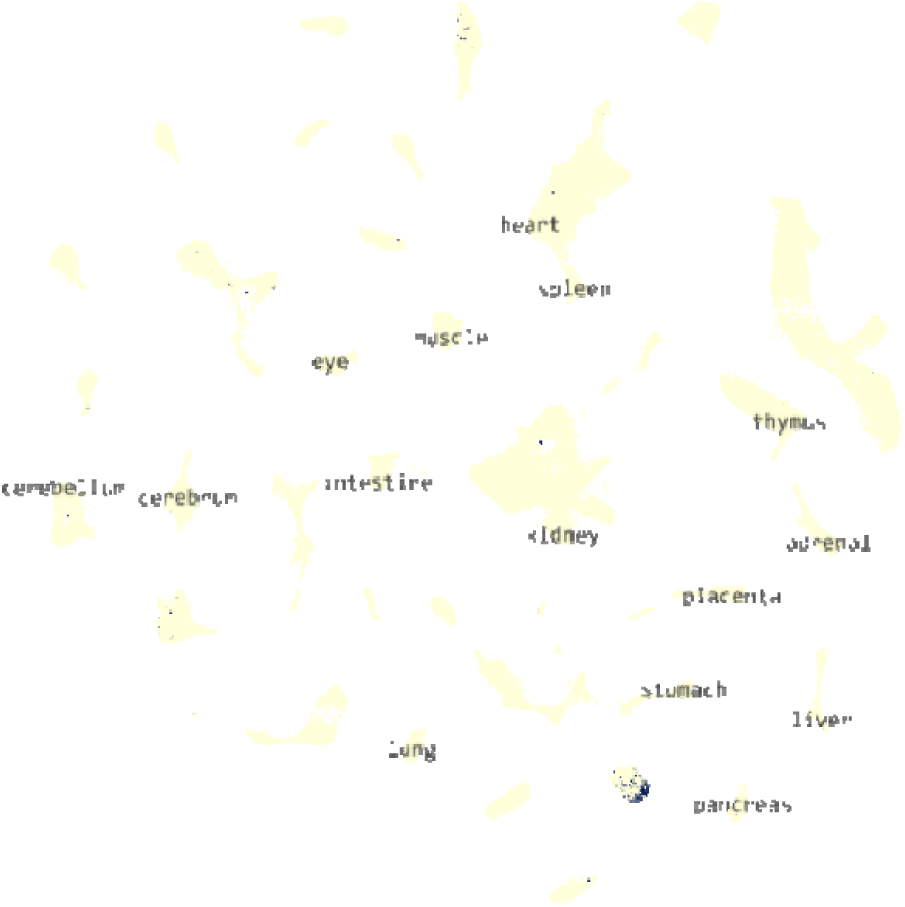
Fetal single cell gene expression expressed in the pancreas for NEC1, where the organs are shown in yellow and the single cells demonstrating expression in navy blue.

**Supplementary figure 22.**
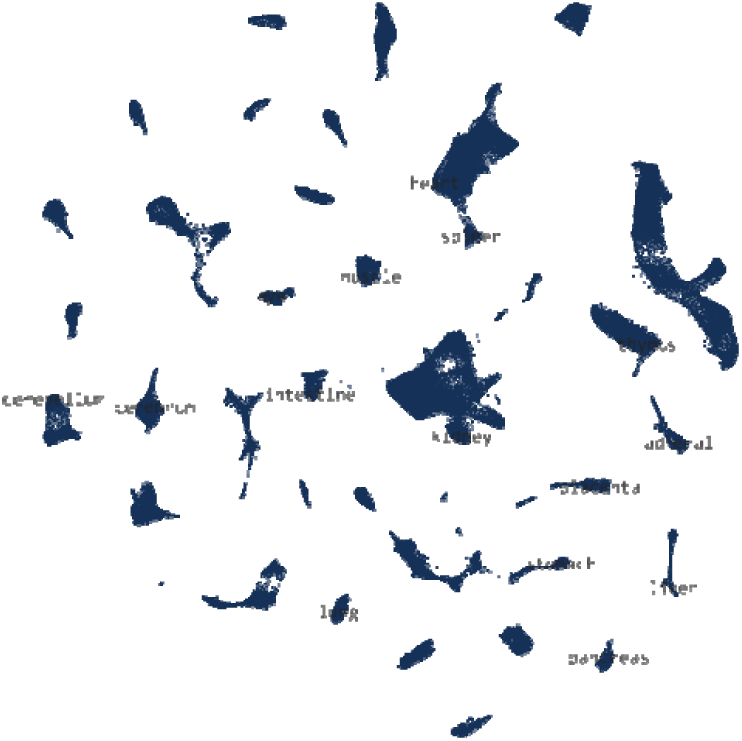
Fetal single cell gene expression expressed tissue-wide for Galectin_4, where the organs are shown in yellow and the single cells demonstrating expression in navy blue.

**Supplementary figure 23.**
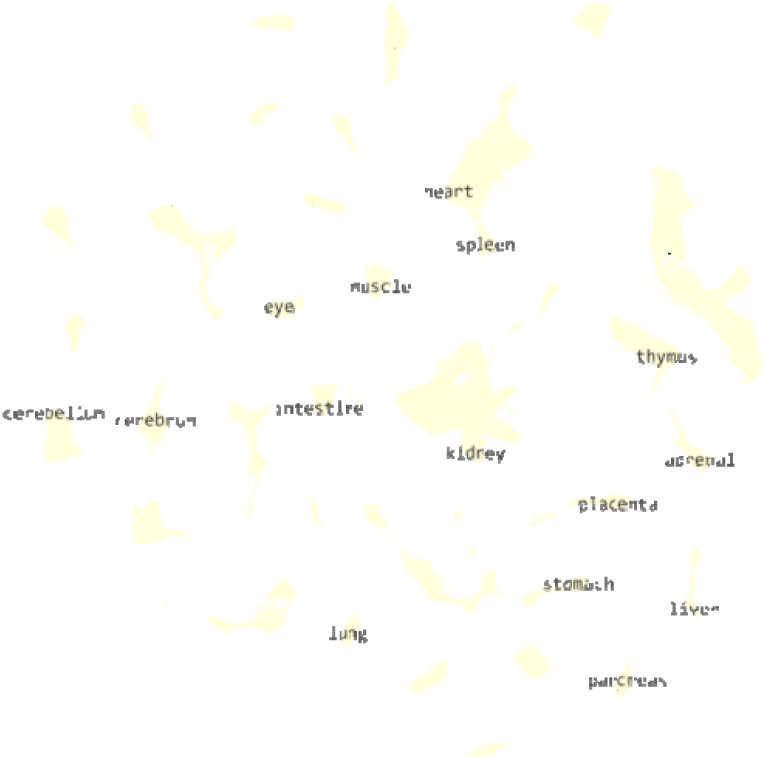
Fetal single cell gene expression expressed tissue-wide for FUT5, where the organs are shown in yellow and the single cells demonstrating expression in navy blue.

**Supplementary figure 24.**
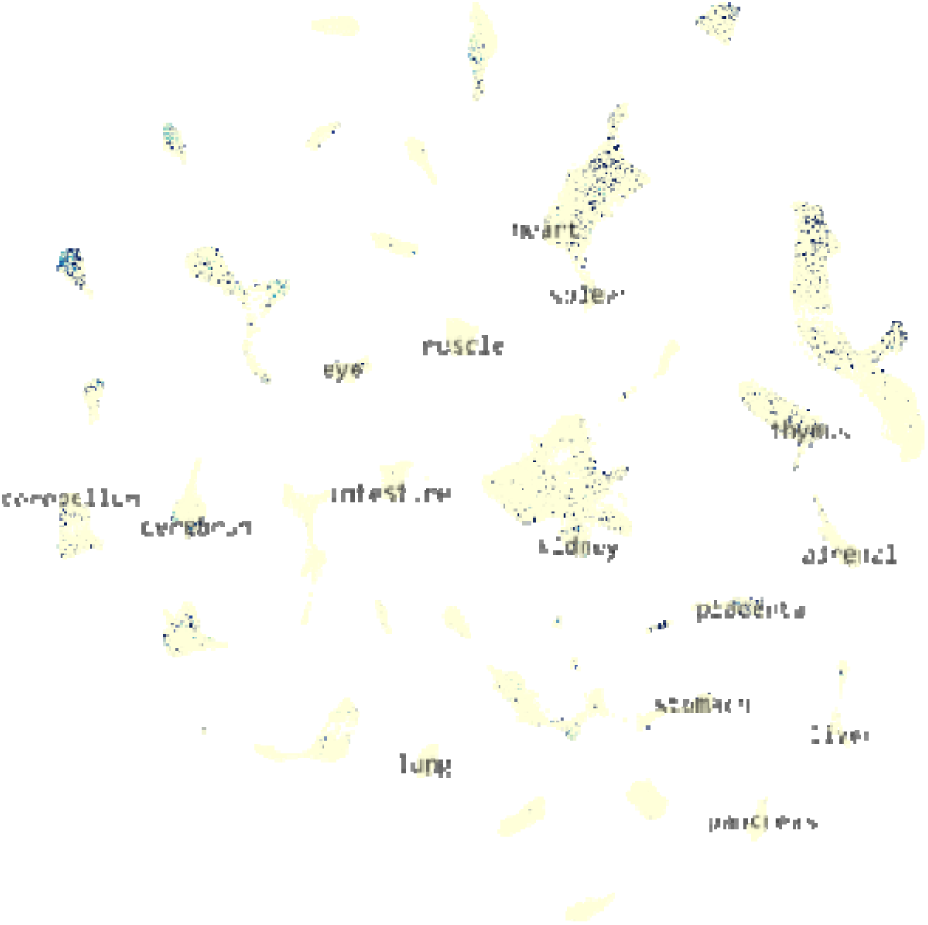
Fetal single cell gene expression expressed tissue-wide for UBS3B, where the organs are shown in yellow and the single cells demonstrating expression in navy blue.

**Supplementary figure 25.**
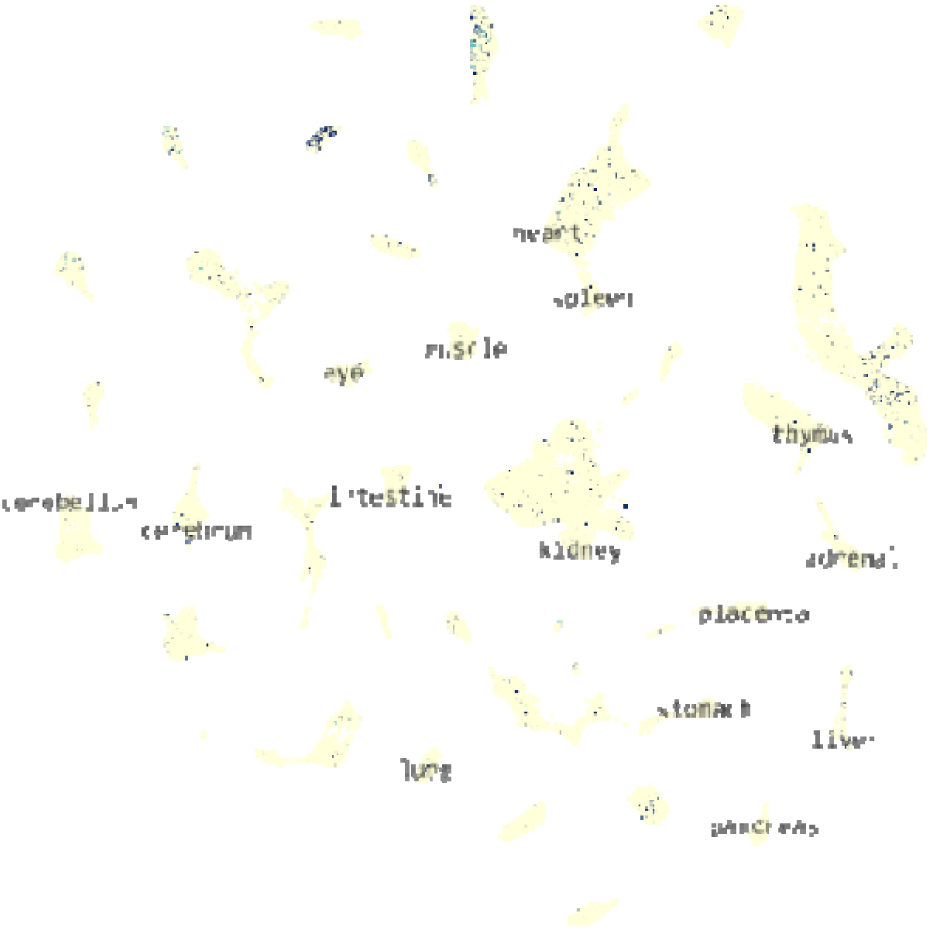
Fetal single cell gene expression expressed tissue-wide for sLeptin_R, where the organs are shown in yellow and the single cells demonstrating expression in navy blue.

**Supplementary figure 26.**
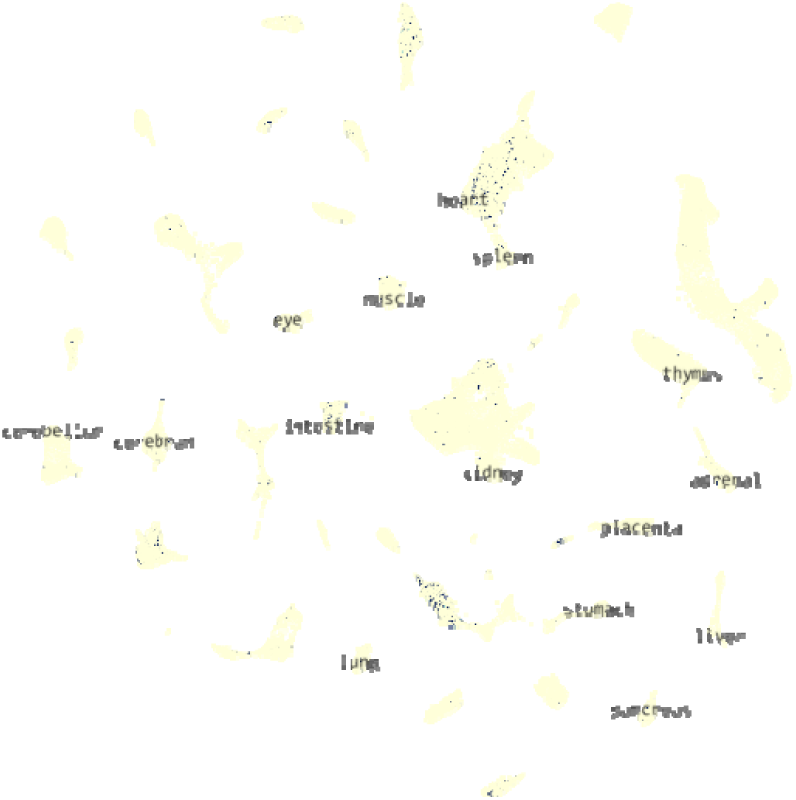
Fetal single cell gene expression expressed tissue-wide for AIF1L, where the organs are shown in yellow and the single cells demonstrating expression in navy blue.

**Supplementary figure 27.**
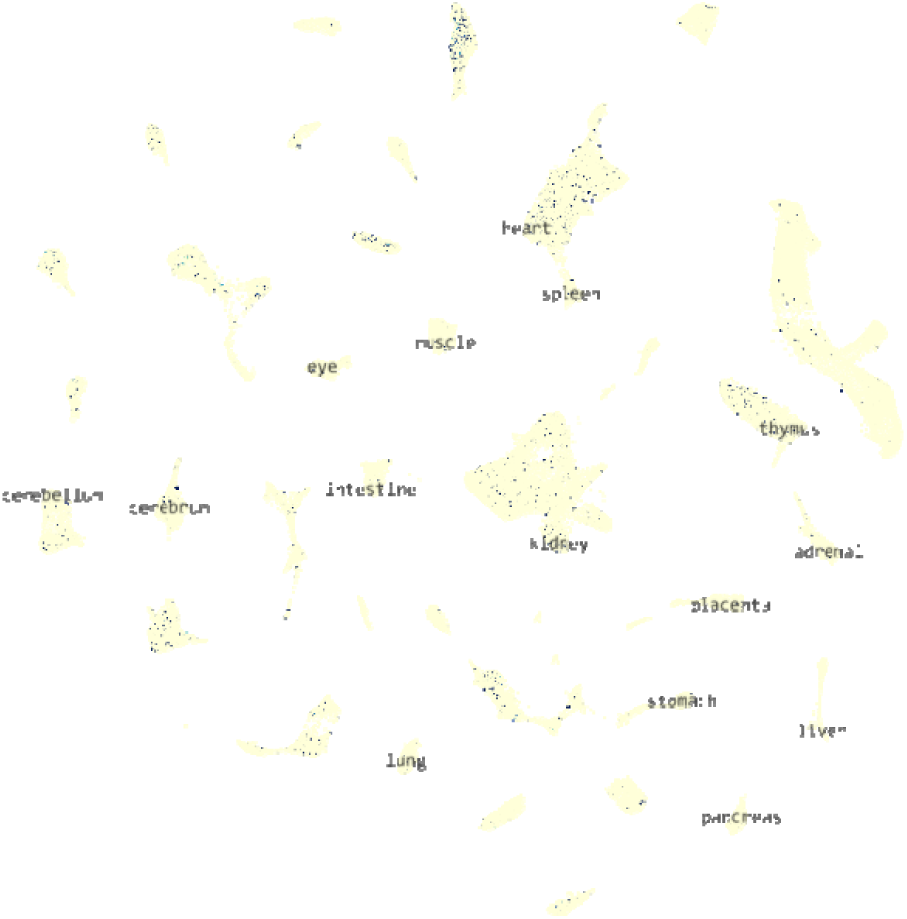
Fetal single cell gene expression expressed tissue-wide for PLCG1, where the organs are shown in yellow and the single cells demonstrating expression in navy blue.

**Supplementary figure 28.**
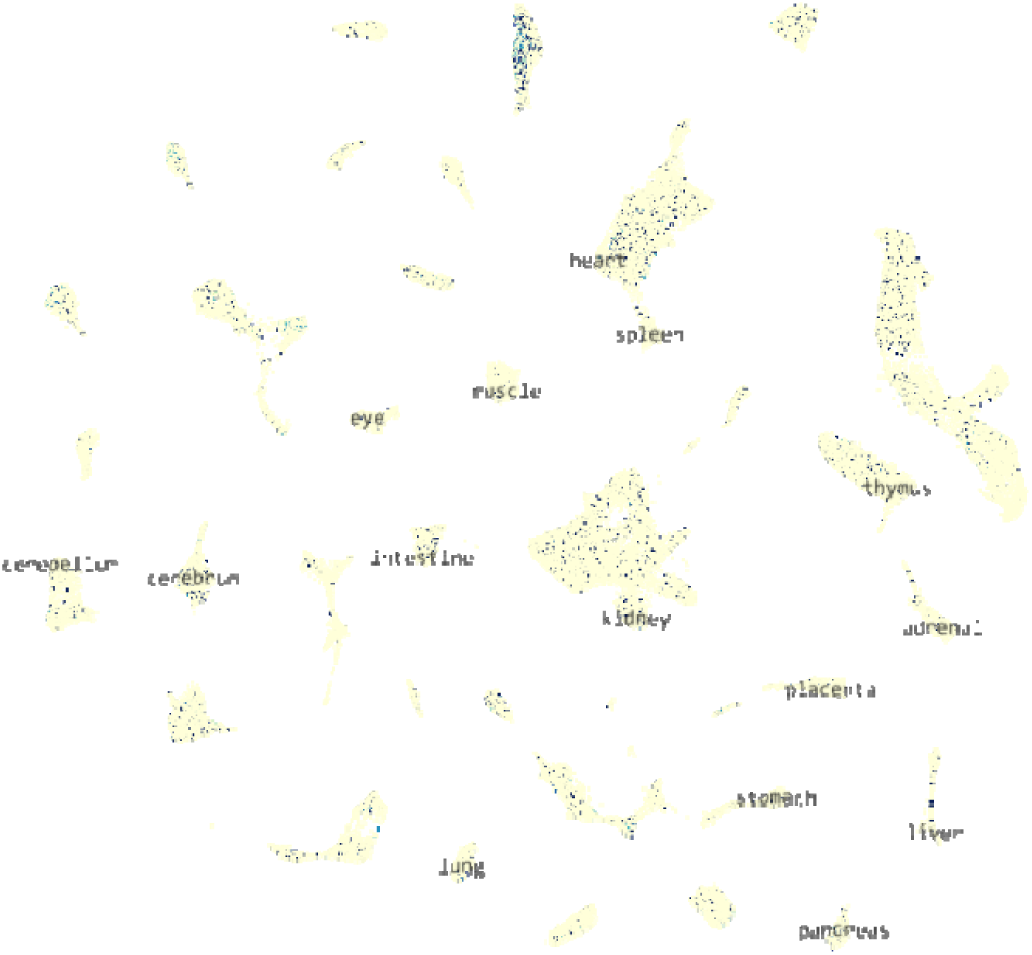
Fetal single cell gene expression expressed tissue-wide for IFN_a_b_R1, where the organs are shown in yellow and the single cells demonstrating expression in navy blue.

**Supplementary figure 29.**
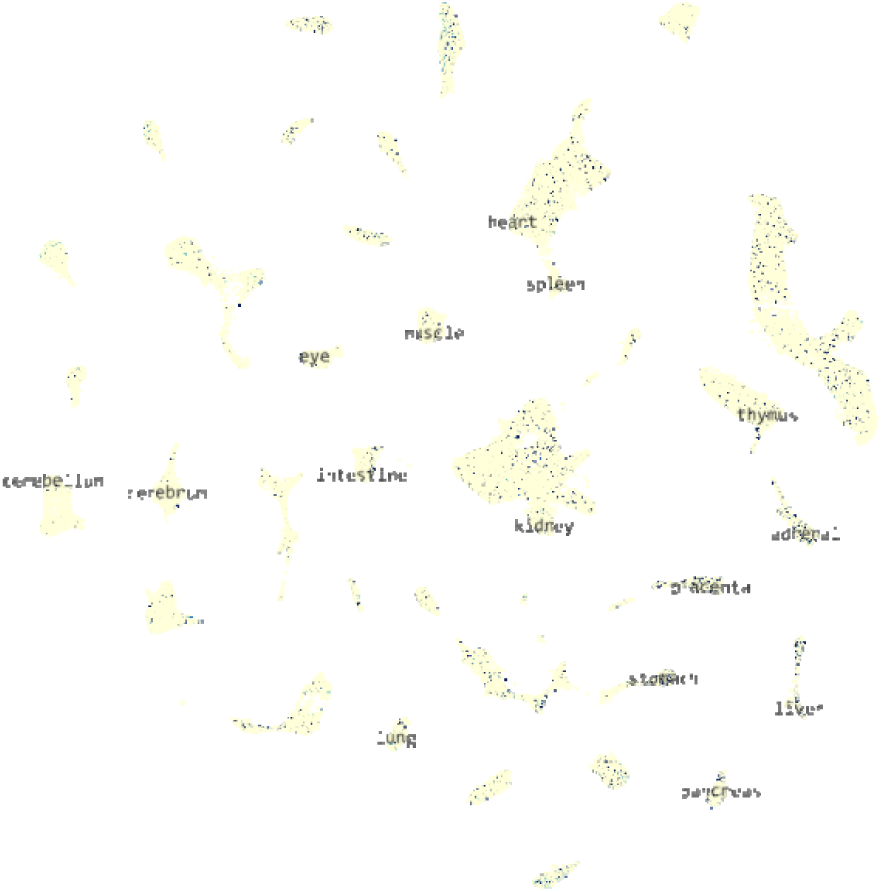
Fetal single cell gene expression expressed tissue-wide for ACADV, where the organs are shown in yellow and the single cells demonstrating expression in navy blue.

**Supplementary figure 22.**
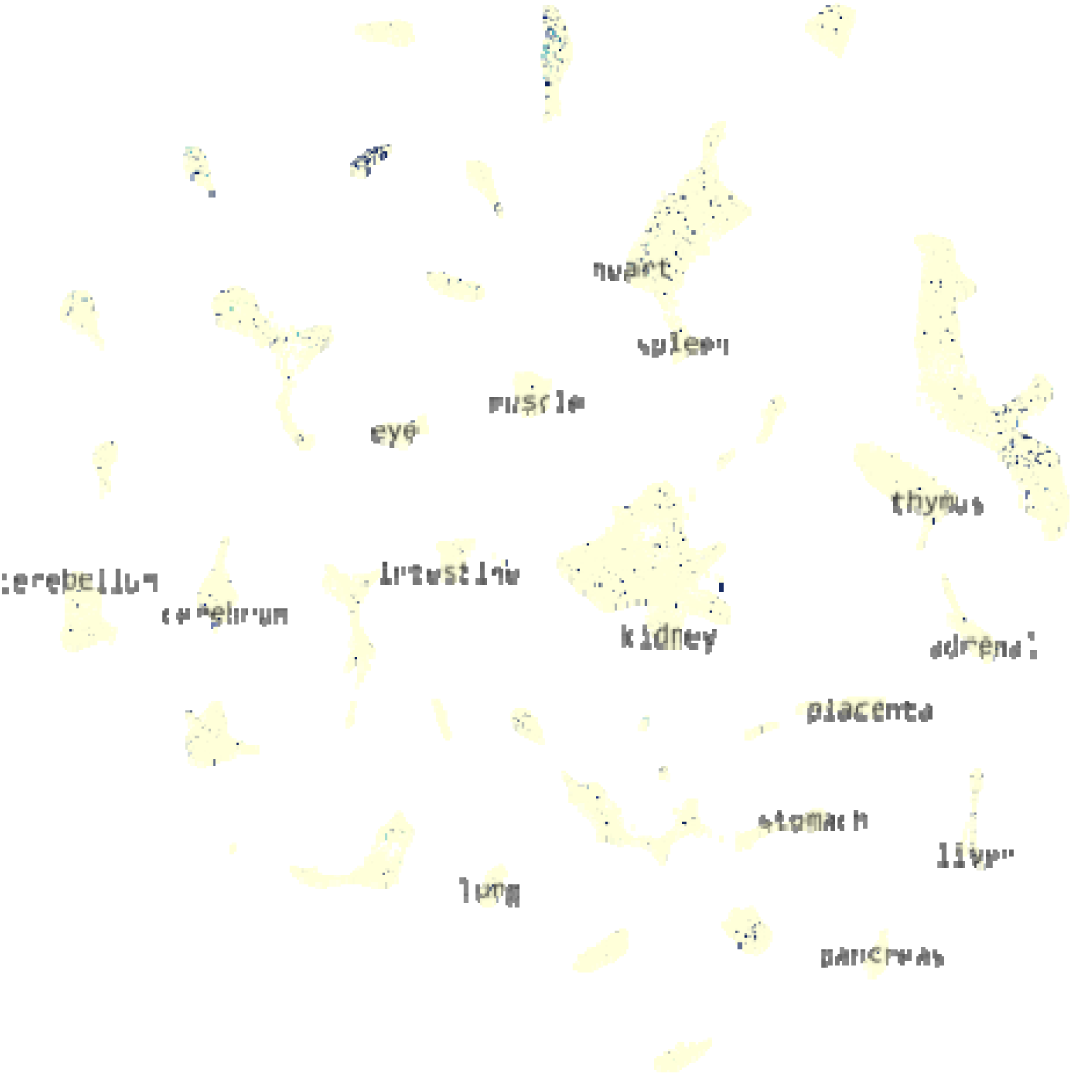
Fetal single cell gene expression expressed tissue-wide for sLeptin_R, where the organs are shown in yellow and the single cells demonstrating expression in navy blue.

## References

1. Küpers LK, Monnereau C, Sharp GC, Yousefi P, Salas LA, Ghantous A, et al. Meta-analysis of epigenome-wide association studies in neonates reveals widespread differential DNA methylation associated with birthweight. Nature Communications. 2019;10(1):1893.

2. Leite DFB, Cecatti JG. Fetal Growth Restriction Prediction: How to Move beyond. The Scientific World Journal. 2019;2019:1519048.

3. Warrington NM, Freathy RM, Neale MC, Evans DM. Using structural equation modelling to jointly estimate maternal and fetal effects on birthweight in the UK Biobank. International Journal of Epidemiology. 2018;47(4):1229–41.

4. Wilcox AJ. On the importance—and the unimportance— of birthweight. International Journal of Epidemiology. 2001;30(6):1233–41.

5. Barry CA-O, Lawlor DA, Shapland CA-O, Sanderson E, Borges MA-O. Using Mendelian Randomisation to Prioritise Candidate Maternal Metabolic Traits Influencing Offspring Birthweight. LID - 10.3390/metabo12060537 [doi] LID - 537. (2218–1989 (Print)).

6. Bond TA, Karhunen V, Wielscher M, Auvinen J, Männikkö M, Keinänen-Kiukaanniemi S, et al. Exploring the role of genetic confounding in the association between maternal and offspring body mass index: evidence from three birth cohorts. International Journal of Epidemiology. 2019;49(1):233–43.

7. Iliodromiti S, Mackay DF, Smith GCS, Pell JP, Sattar N, Lawlor DA, et al. Customised and Noncustomised Birth Weight Centiles and Prediction of Stillbirth and Infant Mortality and Morbidity: A Cohort Study of 979,912 Term Singleton Pregnancies in Scotland. PLOS Medicine. 2017;14(1):e1002228.

8. Warrington NM, Beaumont RN, Horikoshi M, Day FR, Helgeland Ø, Laurin C, et al. Maternal and fetal genetic effects on birth weight and their relevance to cardio-metabolic risk factors. Nature genetics. 2019;51(5):804–14.

9. Beaumont RN, Warrington NM, Cavadino A, Tyrrell J, Nodzenski M, Horikoshi M, et al. Genome-wide association study of offspring birth weight in 861577 women identifies five novel loci and highlights maternal genetic effects that are independent of fetal genetics. (1460–2083 (Electronic)).

10. Ferkingstad E, Sulem P, Atlason BA, Sveinbjornsson G, Magnusson MI, Styrmisdottir EL, et al. Large-scale integration of the plasma proteome with genetics and disease. Nature Genetics. 2021;53(12):1712–21.

11. Zheng J, Haberland V, Baird D, Walker V, Haycock PC, Hurle MR, et al. Phenome-wide Mendelian randomization mapping the influence of the plasma proteome on complex diseases. Nature Genetics. 2020;52(10):1122–31.

12. Zhang G, Bacelis J, Lengyel C, Teramo K, Hallman M, Helgeland Ø, et al. Assessing the Causal Relationship of Maternal Height on Birth Size and Gestational Age at Birth: A Mendelian Randomization Analysis. (1549–1676 (Electronic)).

13. Tyrrell J, Richmond RC, Palmer TM, Feenstra B, Rangarajan J, Metrustry S, et al. Genetic Evidence for Causal Relationships Between Maternal Obesity-Related Traits and Birth Weight. (1538–3598 (Electronic)).

14. Ardissino MA-O, Slob EA-O, Millar O, Reddy RA-OX, Lazzari L, Patel KHK, et al. Maternal Hypertension Increases Risk of Preeclampsia and Low Fetal Birthweight: Genetic Evidence From a Mendelian Randomization Study. (1524–4563 (Electronic)).

15. Brand JA-O, Gaillard RA-O, West JA-O, McEachan RA-O, Wright JA-O, Voerman E, et al. Associations of maternal quitting, reducing, and continuing smoking during pregnancy with longitudinal fetal growth: Findings from Mendelian randomization and parental negative control studies. (1549–1676 (Electronic)).

16. Thompson WA-O, Beaumont RA-O, Kuang AA-O, Warrington NA-OX, Ji Y, Tyrrell JA-O, et al. Higher maternal adiposity reduces offspring birthweight if associated with a metabolically favourable profile. (1432–0428 (Electronic)).

17. Allen NE, Sudlow C, Peakman T, Collins R, Biobank UK. UK biobank data: come and get it. Sci Transl Med. 6. United States2014. p. 224ed4.

18. Haugan A, Birke C, Alsaker E, Høiseth G, Knudsen GP, Tambs K, et al. Cohort Profile Update: The Norwegian Mother and Child Cohort Study (MoBa). International Journal of Epidemiology. 2016;45(2):382–8.

19. Paltiel L, Anita H, Skjerden T, Harbak K, Bækken S, Nina Kristin S, et al. The biobank of the Norwegian Mother and Child Cohort Study – present status. Norsk Epidemiologi. 2014;24(1-2).

20. Corfield EC, Frei O, Shadrin AA, Rahman Z, Lin A, Athanasiu L, et al. The Norwegian Mother, Father, and Child cohort study (MoBa) genotyping data resource: MoBaPsychGen pipeline v.1. bioRxiv. 2022:2022.06.23.496289.

21. McCarthy S, Das S, Kretzschmar W, Delaneau O, Wood AR, Teumer A, et al. A reference panel of 64,976 haplotypes for genotype imputation. Nature Genetics. 2016;48(10):1279–83.

22. Mbatchou J, Barnard L, Backman J, Marcketta A, Kosmicki JA, Ziyatdinov A, et al. Computationally efficient whole-genome regression for quantitative and binary traits. Nature Genetics. 2021;53(7):1097–103.

23. Bulik-Sullivan BK, Loh P-R, Finucane HK, Ripke S, Yang J, Patterson N, et al. LD Score regression distinguishes confounding from polygenicity in genome-wide association studies. Nature Genetics. 2015;47(3):291–5.

24. Wallace C. Statistical Testing of Shared Genetic Control for Potentially Related Traits. Genetic Epidemiology. 2013;37(8):802–13.

25. Liu B, Gloudemans MJ, Rao AS, Ingelsson E, Montgomery SB. Abundant associations with gene expression complicate GWAS follow-up. Nature Genetics. 2019;51(5):768–9.

26. Cao J, O’Day DR, Pliner HA, Kingsley PD, Deng M, Daza RM, et al. A human cell atlas of fetal gene expression. Science. 2020;370(6518):eaba7721.

27. Szklarczyk D, Franceschini A, Wyder S, Forslund K, Heller D, Huerta-Cepas J, et al. STRING v10: protein–protein interaction networks, integrated over the tree of life. Nucleic Acids Research. 2014;43(D1):D447–D52.

28. Benzinou M, Creemers Jw Fau - Choquet H, Choquet H Fau - Lobbens S, Lobbens S Fau - Dina C, Dina C Fau - Durand E, Durand E Fau - Guerardel A, et al. Common nonsynonymous variants in PCSK1 confer risk of obesity. (1546–1718 (Electronic)).

29. Stijnen P, Ramos-Molina B, O’Rahilly S, Creemers JWM. PCSK1 Mutations and Human Endocrinopathies: From Obesity to Gastrointestinal Disorders. Endocrine Reviews. 2016;37(4):347–71.

30. Wei X, Ma X, Lu R, Bai G, Zhang J, Deng R, et al. Genetic Variants in PCSK1 Gene Are Associated with the Risk of Coronary Artery Disease in Type 2 Diabetes in a Chinese Han Population: A Case Control Study. PLOS ONE. 2014;9(1):e87168.

31. Gjesing AP, Vestmar MA, Jørgensen T, Heni M, Holst JJ, Witte DR, et al. The Effect of PCSK1 Variants on Waist, Waist-Hip Ratio and Glucose Metabolism Is Modified by Sex and Glucose Tolerance Status. PLOS ONE. 2011;6(9):e23907.

32. Nead KT, Li A, Wehner MR, Neupane B, Gustafsson S, Butterworth A, et al. Contribution of common non-synonymous variants in PCSK1 to body mass index variation and risk of obesity: a systematic review and meta-analysis with evidence from up to 331 175 individuals. (1460–2083 (Electronic)).

33. Ryan DH. Next Generation Antiobesity Medications: Setmelanotide, Semaglutide, Tirzepatide and Bimagrumab: What do They Mean for Clinical Practice? J Obes Metab Syndr. 2021;30(3):196–208.

34. Blois SM, Dveksler G, Vasta GR, Freitag N, Blanchard V, Barrientos G. Pregnancy Galectinology: Insights Into a Complex Network of Glycan Binding Proteins. (1664–3224 (Electronic)).

35. Schrader S, Unverdorben L, Hutter S, Knabl J, Schmoeckel E, Meister S, et al. Overexpression of galectin-4 in placentas of women with gestational diabetes. (1872–7603 (Electronic)).

36. Ajayi F, Kongoasa N, Gaffey T, Asmann YW, Watson WJ, Baldi A, et al. Elevated expression of serine protease HtrA1 in preeclampsia and its role in trophoblast cell migration and invasion. Am J Obstet Gynecol. 2008;199(5):557.e1–10.

37. Vukojević K, Šoljić V, Martinović V, Raguž F, Filipović N. The Ubiquitin-Associated and SH3 Domain-Containing Proteins (UBASH3) Family in Mammalian Development and Immune Response. Int J Mol Sci. 2024;25(3).

38. Pena LD, van Calcar SC, Hansen J, Edick MJ, Walsh Vockley C, Leslie N, et al. Outcomes and genotype-phenotype correlations in 52 individuals with VLCAD deficiency diagnosed by NBS and enrolled in the IBEM-IS database. Mol Genet Metab. 2016;118(4):272–81.

39. Hoggard N, Hunter L, Duncan JS, Williams LM, Trayhurn P, Mercer JG. Leptin and leptin receptor mRNA and protein expression in the murine fetus and&#x2009;placenta. Proceedings of the National Academy of Sciences. 1997;94(20):11073–8.

40. Kilpeläinen TO, Carli JFM, Skowronski AA, Sun Q, Kriebel J, Feitosa MF, et al. Genome-wide meta-analysis uncovers novel loci influencing circulating leptin levels. Nature Communications. 2016;7(1):10494.

41. Agrawal S, Cerdeira AS, Redman C, Vatish M. Meta-Analysis and Systematic Review to Assess the Role of Soluble FMS-Like Tyrosine Kinase-1 and Placenta Growth Factor Ratio in Prediction of Preeclampsia: The SaPPPhirE Study. (1524–4563 (Electronic)).

42. Andrikos AA-OX, Andrikos D, Schmidt B, Birdir C, Kimmig R, Gellhaus A, et al. Course of the sFlt-1/PlGF ratio in fetal growth restriction and correlation with biometric measurements, feto-maternal Doppler parameters and time to delivery. (1432–0711 (Electronic)).

43. Chang Y-S, Chen C-N, Jeng S-F, Su Y-N, Chen C-Y, Chou H-C, et al. The sFlt-1/PlGF ratio as a predictor for poor pregnancy and neonatal outcomes. Pediatrics & Neonatology. 2017;58(6):529–33.

44. Suhre K, McCarthy MI, Schwenk JM. Genetics meets proteomics: perspectives for large population-based studies. Nature Reviews Genetics. 2021;22(1):19–37.

45. Fauman EB, Hyde C. An optimal variant to gene distance window derived from an empirical definition of cis and trans protein QTLs. BMC Bioinformatics. 2022;23(1):169.

46. Zhao J, Stewart ID, Baird D, Mason D, Wright J, Zheng J, et al. Causal effects of maternal circulating amino acids on offspring birthweight: a Mendelian randomisation study. (2352–3964 (Electronic)).

