## Supplemental Information for "Effects of the maternal and fetal proteome on birth weight: a Mendelian randomization analysis"

Supplementary section

### Methods

#### Discovery - Early Growth Genetics Consortium and UK BioBank

The EGG consortium consists of studies with birth cohort designs, where parents were recruited in pregnancy and birth weight was recorded. Across these cohorts, birth weight data were generally abstracted from medical records or self-reported as adults. The UKBB is a cohort of British residents (aged 37-73 years) recruited from 22 centres across the UK. Birth weight data were recalled and self-reported. Sex of the offspring was not asked.

Genotyping

Data from the EGG consortium were imputed up to the 1000 Genomes Project (Phase 1 v3) reference panel and data from the UK Biobank were imputed up to the HRC reference panel. SNPs were excluded if they were detected in less than 50% of the sample, had an INFO score <0.3, minor allele frequency (MAF) <1%, or the SNPs had not been imputed by HRC. Whilst the analysis was restricted to Europeans, principal components analysis was used to exclude ancestry outliers.

Fixed effects meta-analysis were run in GWAMA (1).

The European-ancestry GWAS meta-analysis of offspring birth weight using the maternal genotypes consisted of three components: i) 12,319 individuals from 10 GWAS in the EGG consortium were imputed using the HapMap 2 reference panel; ii) 7,542 individuals from two GWAS in the EGG consortium were imputed using the HRC panel; and iii) 190,406 individuals of White European origin from the UKBB. In contrast, the European-ancestry GWAS meta-analysis of offspring birth weight using the fetal genotypes consisted of two parts: i) 80,745 individuals from 35 studies in the EGG Consortium from Europe, the United States and Australia; and ii) 217,397 individuals from the UKBB. Variant filtering criteria are detailed in the **Supplementary information**. A fixed-effects meta-analysis of the EGG and UKBB data using the maternal genetics was conducted using GWAMA (N = 210,267).

#### Replication - The Norwegian Mother Father and Child birth cohort

For our replication analysis, we performed GWAS using the Norwegian Mother Father and Child birth cohort (MoBa). The Medical Birth Registry of Norway, established in 1967, is a national health registry containing information about all births in Norway. MoBa has been linked to the Medical Birth Registry of Norway using unique personal identification numbers. In MoBa, plasma blood samples were obtained from both parents during pregnancy and from mothers and children (umbilical cord) at birth. Genotyping, pre-imputation quality control, and imputation procedures were described in detail elsewhere. There were five projects contributed to MoBa genetics 1.0 (<https://github.com/folkehelseinstituttet/mobagen>). We have used the latest data release here, quality control undertaken by the MoBaPsychGen pipeline (2). Detailed elsewhere, the MoBaPsychGen pipeline, was developed to account for the complex structure of the MoBa genotype data. The pipeline includes QC on both single nucleotide polymorphism (SNP) and individual level. Phasing and imputation were performed using the publicly available Haplotype Reference Consortium release 1.1 panel as a reference. Phenotypic information from the MBRN and MoBa questionnaires were used to identify biological sex, year of birth, reported parent-offspring (PO) relationships, and multiple births (only available in the offspring). The pipeline consists of best-guess genotypes data on 207,409 MoBa participants for 6,981,748 variants. We conducted analyses separately for mothers and children.

###### Genotyping

In total, 238,001 MoBa samples were sent to be genotyped in 24 genotyping batches with varying selection criteria, genotyping arrays, and genotyping centres. Full details about genotyping of MoBa are described in the pipeline paper (2). Scripts used throughout the MoBaPsychGen pipeline are available on GitHub: <https://github.com/psychgen/MoBaPsychGen-QC-pipeline>. They used the HRC reference panel with SHAPEIT2 and duoHMM and HRC and IMPUTE4 for imputation.

Dosages were converted to best-guess hard-call genotype data using a certainty threshold of 0.7. Post imputation QC undertook the following steps: removing SNPs with - (1) imputation quality (INFO) score = 0.8; (2) MAF < 1%; (3) call rate < 95%; (4) HWE p-value < 1.00*10-6; (5) discordant (concordance rate < 97%) in true duplicates (expected duplicates confirmed by genetic data PI_HAT > 0.95); (6) > 1% ME (with remaining ME set to missing); and (7) association with genotype batch at a p-value < 5x10^-8^.

In terms of individuals, they were excluded if they had - call rate < 98%; (2) ± 3 standard deviations from the mean heterozygosity across all individuals; (3) the individual from each true duplicate pair with the lowest call rate; (4) all relatedness checks described in module 2; (5) cryptic relatedness; (6) ME > 5% in families; (7) ancestry outliers (PCA MoBa & 1000 Genomes); and (8) subpopulation outliers (PCA MoBa).

The pipeline consists of best-guess genotypes data on 207,409 MoBa participants for 6,981,748 SNPs.

###### Genome Wide Association Study

GWAS were run in Rversion 4.1.0 using regenie software (v 3.1.2) (3). In order to run the GWAS, we removed duplicated phenotypic IDs mapping to unique genetic IDs (87,651 mothers and 82,639 children) and individuals with missing phenotype information.

To carry out step 1 of regenie, we used a pre-defined random set of 455,827 imputed SNPs. We began with a list of 491,637 variants which were directly genotyped in >=1 imputation batch and had imputation quality INFO score >0.99785. We then removed 35,810 variants in the major histocompatibility complex (MHC) region located on chromosome 6 (as a precaution due to reports of apparent inter-chromosomal linkage disequilibrium/mis-mapping affecting variants in the MHC region [<https://doi.org/10.1101/2020.08.03.235150>, <https://doi.org/10.1038/s41588-021-00870-7>]).

The method then uses a second ridge regression (referred to as Level 1 by regenie) to combine the M predictors (M = 5*491,637/1,000 = 2,459) into a single predictor, which is then decomposed into 22 Leave One Chromosome Out (LOCO) predictions, using linear or logistic regression for continuous or binary phenotypes, respectively. The resulting LOCO predictions are then used as a covariate in step 2 when imputed SNPs on the relevant chromosome are tested. All of the predictions at Level 0 and Level 1 are obtained within a cross-validation (CV) scheme to prevent over-fitting.

In step 1 of regenie, we fit a whole genome regression model to capture the phenotypic variance attributable to genetic effects captured by a subset of genetic markers. The resulting model fit was then used to predict birth weight, that will be used as covariates in step 2 to account for population structure and relatedness. In step 2, all 6,981,748 imputed variants on the same set of individuals were tested for association with birth weight, conditional on the predictions from step 1 and covariates. These variants were read in blocks of B=200 markers, and tested for association, to avoid having all markers stored in memory at once. The summary statistics data was then transferred from the MoBa servers to ours.

Mutual adjustment

As detailed in previous work, the correlation between maternal, fetal and maternal genotype can confound results. We undertook an adjustment in the MoBa summary statistics to account for this. We used the summary statistics from the birth weight GWAS In both mothers and offspring. We ran LD score regression (ldsc) to obtain genetic covariance intercepts between mother and child. These were used as inputs in to DONUTS (DecOmposing Nature and nUrTure using GWAS Summary statistics) (4). DONUTS uses these to estimate direct and indirect genetic effects at the SNP level, and accounts for GWAS sample overlap and assortative mating. The output of this was then used for our two sample MR replication analyses in both maternal and fetal genotype.

References

1.	Mägi R, Morris AP. GWAMA: software for genome-wide association meta-analysis. BMC Bioinformatics. 2010;11(1):288.

2.	Corfield EC, Frei O, Shadrin AA, Rahman Z, Lin A, Athanasiu L, et al. The Norwegian Mother, Father, and Child cohort study (MoBa) genotyping data resource: MoBaPsychGen pipeline v.1. bioRxiv. 2022:2022.06.23.496289.

3.	Mbatchou J, Barnard L, Backman J, Marcketta A, Kosmicki JA, Ziyatdinov A, et al. Computationally efficient whole-genome regression for quantitative and binary traits. Nature Genetics. 2021;53(7):1097-103.

4.	Wu Y, Zhong X, Lin Y, Zhao Z, Chen J, Zheng B, et al. Estimating genetic nurture with summary statistics of multigenerational genome-wide association studies. Proceedings of the National Academy of Sciences. 2021;118(25):e2023184118.
